# Temporal evolution of multiday, epileptic functional networks prior to seizure occurrence

**DOI:** 10.1101/2022.08.10.22278581

**Authors:** Petroula Laiou, Andrea Biondi, Elisa Bruno, Pedro Viana, Joel S Winston, Zulqarnain Rashid, Yatharth Ranjan, Pauline Conde, Callum Stewart, Shaoxiong Sun, Yuezhou Zhang, Amos Folarin, Richard JB Dobson, Andreas Schulze-Bonhage, Matthias Duempelmann, Mark P Richardson, RADAR-Consortium

**Affiliations:** Department of Biostatistics and Health Informatics, Institute of Psychiatry, Psychology and Neuroscience, King’s College London, United Kingdom; Department of Basic and Clinical Neuroscience, Institute of Psychiatry, Psychology and Neuroscience, King’s College London, United Kingdom; Faculty of Medicine, University of Lisbon, Lisbon, Portugal; Institute of Health Informatics, University College London, London, United Kingdom; NIHR Biomedical Research Centre at South London and Maudsley NHS Foundation Trust and King’s College London, London, United Kingdom; Health Data Research UK London, University College London, London, United Kingdom; NIHR Biomedical Research Centre at University College London Hospitals NHS Foundation Trust, London, United Kingdom; Epilepsy Center, Medical Center-University of Freiburg, Faculty of Medicine, University of Freiburg, Freiburg, Germany

**Keywords:** epilepsy, functional network, evolving network, graph theory, EEG, ECG, seizure lateralization

## Abstract

Epilepsy is one of the most common neurological disorders, characterized by the occurrence of repeated seizures. Given that epilepsy is considered a network disorder, tools derived from network neuroscience may confer the valuable ability to quantify properties of epileptic brain networks. In this study we use well-established brain network metrics (i.e., mean strength, variance of strength, eigenvector centrality, betweenness centrality) to characterize the temporal evolution of epileptic functional networks over several days prior to seizure occurrence. We infer the networks using long-term electroencephalographic recordings from 12 people with epilepsy. We found that brain network metrics are variable across days and show a circadian periodicity. In addition, we found that in 9 out of 12 patients the distribution of variance of strength in the day (or even two last days) prior to seizure occurrence is significantly different compared to the corresponding distributions on all previous days. Our results suggest that brain network metrics computed from EEG recordings could potentially be used to characterize brain network changes that occur prior to seizures, and ultimately contribute to seizure warning systems.

## 1. Introduction

The human brain is a dynamical system that undergoes several dynamic changes due to, for example, different cognitive processes, states of vigilance or motor tasks [McKenna_1994]. The brains of people with epilepsy (PWE) have an additional dynamic change in their dynamic repertoire, making transitions between “normal activity” and “seizure activity” [Da Silva_2003]. The occurrence of repeated seizures is the main characteristic of epilepsy and seizures that cannot be controlled by medications cause significant burden in the lives of PWE [Fisher_2000, Dumanis_2017]. Epilepsy is considered to be a network disorder, with several studies suggesting that even focal seizures arise from large-scale networks [Spencer_2002, Richardson_2012, Besson_2017, Bartolomei_2017]. A network can be characterized as being composed of nodes which are connected with edges. In a large-scale brain network, the nodes represent brain regions whilst the edges denote statistical interactions between the nodes (functional network) or anatomical connections between the nodes (structural network) [Sporns_2018]. Hence, to better understand epilepsy it may be valuable to study the topological properties of the epileptic brain networks.

Brain networks can be studied at different spatial and temporal scales using different data modalities [Bassett_2018]. Several studies that aimed to investigate the temporal evolution of epileptic brain networks from minutes to hours and days used intracranial electroencephalographic recordings (iEEG) and built functional networks [Kramer_2008, Schindler_2008, Kuhnert_2010, Lehnertz_2014, Geier_2017]. They assumed each iEEG channel constituted one node and inferred statistical connections between the nodes using linear and non-linear connectivity measures. To characterize topological properties of the brain networks they used metrics from graph theory such as degree (number of links of nodes), betweenness centrality (quantifying the influence that a node has over the flow of information in the network), clustering coefficient (quantifying the tendency of nodes to cluster together) and shortest path length [Newman_2003, Rubinov_2010]. These studies reported high temporal variability of the graph theory metrics across many days with the presence of daily rhythms having a key role in this temporal variation. Moreover, they reported that the brain network evolves from a random structure to a more regular structure during seizures with increasing randomness after seizure termination.

Although iEEG recordings have excellent data quality, their acquisition is highly invasive as it requires brain surgery. In addition, the iEEG electrodes are implanted in spatially limited brain regions and hence do not record electrical activity from the whole brain. In this study, we use longitudinal non-invasive scalp electroencephalographic recordings (EEG) from PWE and build evolving functional networks that span from 2 to 11 days. Using metrics from graph theory we quantify the topological properties of the brain networks and explore their temporal evolution across many days prior to seizure occurrence. We further investigate the presence of periodic patterns in this temporal evolution. Considering the influence of daily rhythms in the temporal evolution of the graph theory metrics, we ask whether the graph metrics that are computed from brain networks of the day prior to seizure occurrence show particular changes in the brain network topology compared to the previous days. Since the EEG acquisition was accompanied with an electrocardiogram (ECG) acquisition, we also investigated the temporal evolution of cardiac metrics (heart rate and heart rate variability). Finally, we discuss the study findings and highlight their importance.

## 2. Materials and Methods

### 2.1. Participants recruitment

Study participants were recruited from July 2017 to February 2020 via the European RADAR-CNS (Remote Assessment of Disease and Relapse - Central Nervous System) study. During this period, 71 participants with an epilepsy diagnosis were admitted to the Epilepsy Monitoring Unit (EMU) of King’s College Hospital, London as part of their clinical workup. The length of the patients’ stay in the EMU was no more than two weeks and was determined by clinical considerations. All patients were monitored continuously via a video-EEG system that recorded simultaneously brain and cardiac activity resulting in 21 channel EEG (Fp1, Fp2, F7, F3, Fz, F4, F8, T3, C3, Cz, C4, T4, T5, A1, A2, P3, Pz, P4, T6, O1, O2; electrode placement according to the modified Maudsley system) and two ECG signals. The sampling frequency was either 256 Hz or 512 Hz. An expert neurologist (E.B.) reviewed all EEG recordings and annotated the timestamps of the seizures onset and offset. More details regarding the data acquisition can be found in Bruno et al. [Bruno_2021].

### 2.2. Inclusion and Exlusion criteria

Since we were interested in the multiday temporal evolution of brain networks prior to seizure occurrence we excluded from the analysis all patients who did not experience seizures during their stay in the EMU. In addition, patients who had seizures for which there was not at least a two-day seizure-free period prior to the seizure occurrence were also excluded from this analysis. Hence, the final dataset that we analyzed comprised 12 participants (one seizure per patient) for whom the seizure-free period prior to the seizure occurrence spanned from 2 to 15 days (Table 1).

**Table 1.** Demographic information of the study participants.

| Patient ID | Sex | Seizure focus | Number of analyzed days prior to seizure occurrence | Number of analyzed EEG epochs | Number of analyzed ECG epochs |
| --- | --- | --- | --- | --- | --- |
| KCL 1 | male | right | 11 | 262 | 258 |
| KCL 2 | female | unclear | 6 | 142 | 128 |
| KCL 3 | female | unclear | 5 | 110 | 0 |
| KCL 4 | male | left | 4 | 91 | 64 |
| KCL 5 | female | left | 3 | 61 | 0 |
| KCL 6 | male | left | 3 | 64 | 41 |
| KCL 7 | male | unclear | 3 | 70 | 69 |
| KCL 8 | female | right | 3 | 71 | 71 |
| KCL 9 | male | unclear | 3 | 70 | 68 |
| KCL 10 | male | unclear | 2 | 45 | 45 |
| KCL 11 | male | unclear | 2 | 47 | 46 |
| KCL 12 | male | unclear | 2 | 48 | 0 |

### 2.3. Ethics statement

The study was conducted in accordance with the Declaration of Helsinki. All participants gave written informed consent and the study procedures were approved by the London Fulham Research Ethics Committee (16/LO/2209; IRAS project ID216316)

### 2.4. EEG measurements

We visually inspected the available EEG recordings, and selected a 20 sec artifact-free epoch from every hour (i.e., 24 EEG epochs/day). Across all patients, the minimum amount of available daily EEG epochs was 21 (missing epochs originated due to the disconnection of the electrodes or low signal quality).

#### 2.4.1. Data preprosessing

An overview of the data preprocessing and analysis pipeline is illustrated in Figure 1. First, we excluded from the analysis the Fp1, Fp2, A1, A2 channels due to the frequent presence of eye/muscle artifacts. Hence, the EEG channels that we analyzed comprised 17 channels. All EEG data were downsampled to 256 Hz, re-referenced to the median and band-passed filtered with a fourth-order Butterworth filter (forward and backward filtering was applied to minimize phase distortions) in four frequency bands, i.e., delta (1-4 Hz), theta (4-8 Hz), alpha (8-13 Hz) and theta (13-25 Hz).

**Figure 1.**
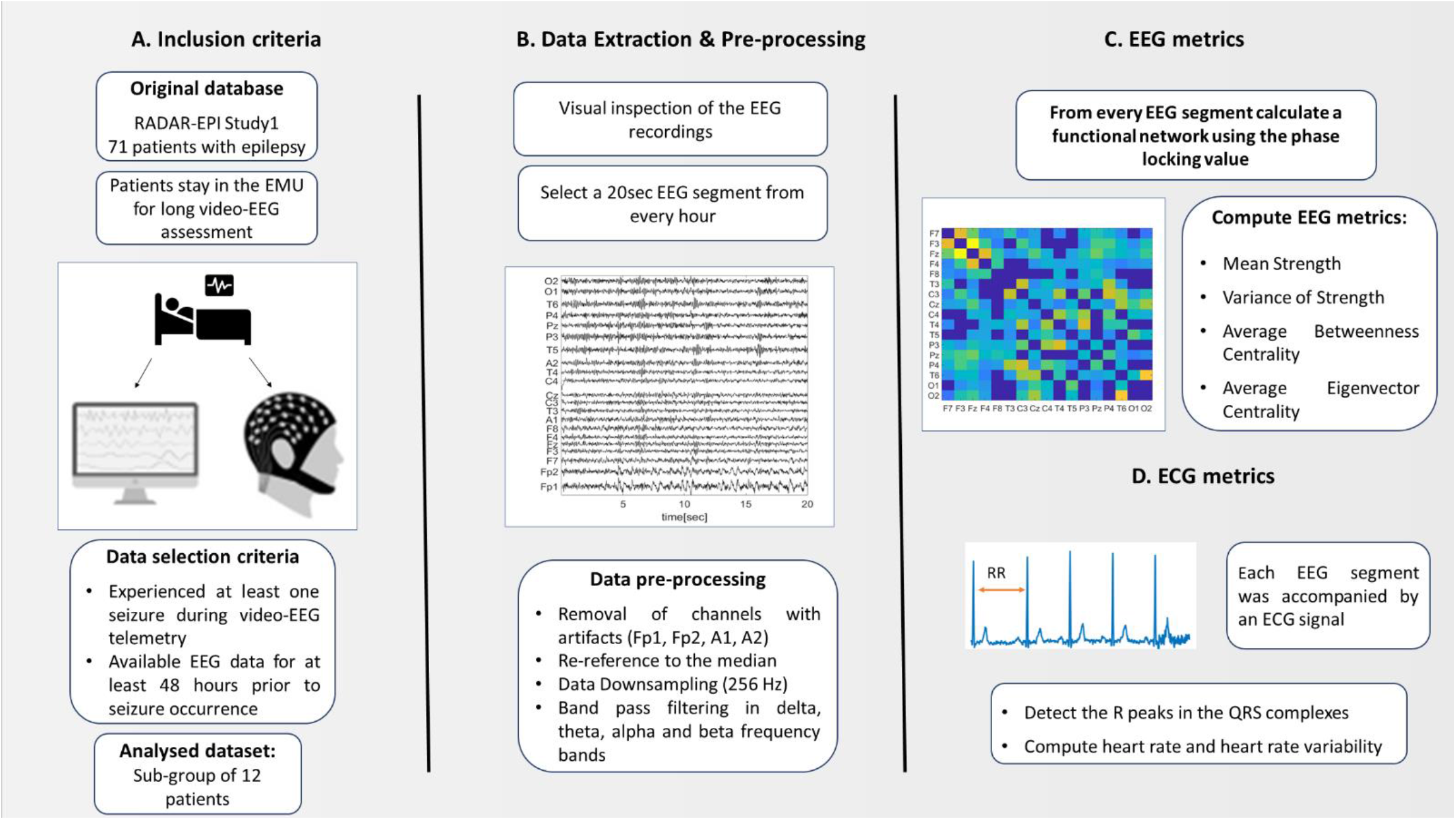
Summary of the data preprocessing and analysis pipeline.

#### 2.4.2. Functional Network

We built functional networks for each 20 sec EEG epoch by associating the network nodes to electrodes, whilst the edges between the nodes *i* and *j* were derived by the phase locking value (PLV) [Tass_1998, Lachaux_1999, Mormann_2000],

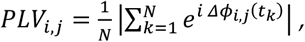

where *N* is the number of samples and *Δϕ*_*i,j*_(*t*_*k*_) is the instantaneous phase difference between the EEG signals that correspond to nodes *i* and *j*. The phases were extracted from the filtered signals using the Hilbert transform. To account for the possible effects of volume conduction [Bastos_2016], connections at zero time lag 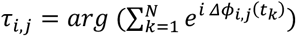 were discarded. Further, to account for the presence of connections that are due to chance or due to the finite length of the signals we additionally used surrogate signals. In particular, for every pair of signals, we generated 99 surrogate pairs of signals using the iterative amplitude-adjusted Fourier transform (IAAFT) with 10 iterations [Schreiber_1996, Schreiber_2000]. Hence, for every PLV value of the (*i, j*) pair of nodes we obtained a distribution of 99 PLV values from the surrogate signals. If the PLV value of the (*i, j*) pair was higher than the 95% of the PLV values of the surrogate pairs, it was retained as a weight between the nodes i and j, otherwise it was set to zero. This process was executed for all four frequency bands and hence for each 20 sec EEG epoch we obtained four functional networks.

#### 2.4.3. EEG metrics

Following the computation of the undirected, weighted functional networks we characterized their structure by using brain network metrics from graph theory such as strength, eigenvector centrality and betweenness centrality [Newman_2003, Rubinov_2010]. The strength of a node is the sum of its weights whilst the betweenness centrality of a node denotes the number of times for which the shortest path lengths of other nodes pass through that node. The eigenvector centrality is a measure of importance of a node. i.e., nodes with large eigenvector centrality are connected to other central nodes. In our study, we are interested in global network properties and thus we computed the mean strength (*S*^*M*^), average betweenness centrality(*C*^*B*^), average eigenvector centrality (*C*^*E*^) which are the average of strength, betweenness and eigenvector centrality across all nodes. In addition, we computed the variance of strength (*S*^*V*^) which is the variance of the strength across all nodes.

### 2.5. ECG metrics

As is clinically standard, the acquisition of the EEG signals was performed simultaneously with an ECG acquisition and hence the selected 20-sec EEG segments were accompanied by two channels of ECG data. Therefore, along with the temporal evolution of the EEG metrics we sought to investigate the temporal evolution of ECG metrics. In each 20 sec epoch from the two available ECG signals, we selected the signal with the best quality. In the case where neither ECG channel showed reasonable quality, we discarded the corresponding ECG epoch from the analysis. If the amount of the available ECG epochs was less than 16 (i.e., 2/3 of the expected amount of daily epochs) we discarded that patient from this analysis. This was the case for 3 out of the 12 analyzed patients (Table 1). Similarly to the EEG acquisition, the sampling frequency of the ECG signal was either 256 Hz or 512 Hz.

#### 2.5.1. HR and HRV metrics

After downsampling the ECG signal to 256Hz, we used the Pan-Tompkins algorithm [Pan_1985] to detect the QRS complexes. Using detected R-peaks of the QRS complexes, we computed the heart rate (HR) and heart rate variability (HRV) for each ECG epoch. We estimated the HR by multiplying by three the number of R-peaks of the corresponding ECG segment (i.e., 3 × 20 *sec* = 1min), whilst the HRV was estimated by the root mean square of the successive difference (RMSSD) of the R-peaks [Shaffer_2017, Shaffer_2020].

### 2.6. Statistical Analysis

For each EEG and ECG metric we obtained a distribution of values whose number was equal to the number of the analysed 20 sec epochs. To account for local, single-sample fluctuations that occur in the distributions of the EEG/ECG measurements, all values were smoothed with a moving average backward filter of lag 5 (i.e. 5 samples) and normalized in the [0, 1] range. In addition, to evaluate whether the distribution of the EEG/ECG metrics of the day before the seizure occurrence was statistically significantly larger or smaller compared to the corresponding distributions of the previous days we performed multiple one-sided non-parametric Wilcoxon rank-sum tests. For instance, if a patient had six seizure-free days before the seizure occurrence, we applied five one-sided Wilcoxon rank-sum tests between the EEG metrics of the day prior to seizure i.e., *d*_−1_ and the EEG metrics of each previous day {*d*_−2_, *d*_−3_, *d*_−4_, *d*_−5_, *d*_−6_}. To decide the side of the non-parametric test (i.e., right or left sided) we compared the medians of the EEG metrics distributions from the day *d*_−1_ and the furthest day from *d*_−1_ i.e., day *d*_−6_. If *median*(*d*_−1_) > *median*(*d*_−6_) we applied one-sided (right tail) tests, otherwise, one-sided (left tail) tests. Results deemed significant for p-values<0.05. To account for the effect of multiple comparisons (number of EEG/ECG metrics, number of frequency bands, number of seizure-free days prior to the seizure occurrence) we applied a Benjamini-Hochberg false discovery rate correction [Benjamini_1995] at a significance level of 5%. In the case for which the EEG/ECG metrics of the day before seizure were statistically significantly higher or lower compared to the corresponding metrics of all the other previous days, we performed a bootstrapping approach to ensure that the results were not due to chance. In particular, we randomly shuffled 100 times the metrics values across different days (i.e., destroying the effect of the day whilst keeping the same number of points in each day) and computed the number of times for which we observed the same pattern (i.e., the metrics of the day before the seizure to be higher or lower compared to the previous days). From this empirical distribution, we then obtained a p-value. Results were deemed significant if *p* − *value* < 0.05. All calculations and data visualization were performed in MATLAB R2020b (MathWorks) and Python version 3.10.0. In addition, we used the Brain Connectivity Toolbox [Rubinov_2010] as well as the Raincloud Plots multi-platform tool [Allen_2019].

## 3. Results

### 3.1. Temporal Evolution of the EEG and ECG metrics

To gain insight into how brain network metrics vary across time and in particular whether they present any particular behavior close to seizure occurrence we started our analysis by illustrating the temporal evolution of the computed EEG metrics. Figure 2 demonstrates the temporal evolution of the brain network metrics (mean strength *S*^*M*^, variance of strength *S*^*V*^, average betweenness centrality *C*^*B*^, average eigenvector centrality *C*^*E*^) in the alpha band, as well as the ECG metrics (HR and HRV) for the two participants (KCL1 and KCL2) who had the longest data available for analysis. We make three observations. First, there is a high variability in both EEG and ECG metrics across time. This variability occurs regardless the time of the day or the period of the seizure occurrence. Second, in the KCL 1 participant, the mean strength *S*^*M*^and variance of strength *S*^*V*^ tend to take larger values in the days closer to seizure occurrence, i.e., days -1, -2 show larger values compared to days -11, -10. Third, all metrics and in particular the ECG metrics, show a partly periodic behavior, that can be possibly attributed to the presence of daily rhythms.

**Figure 2.**
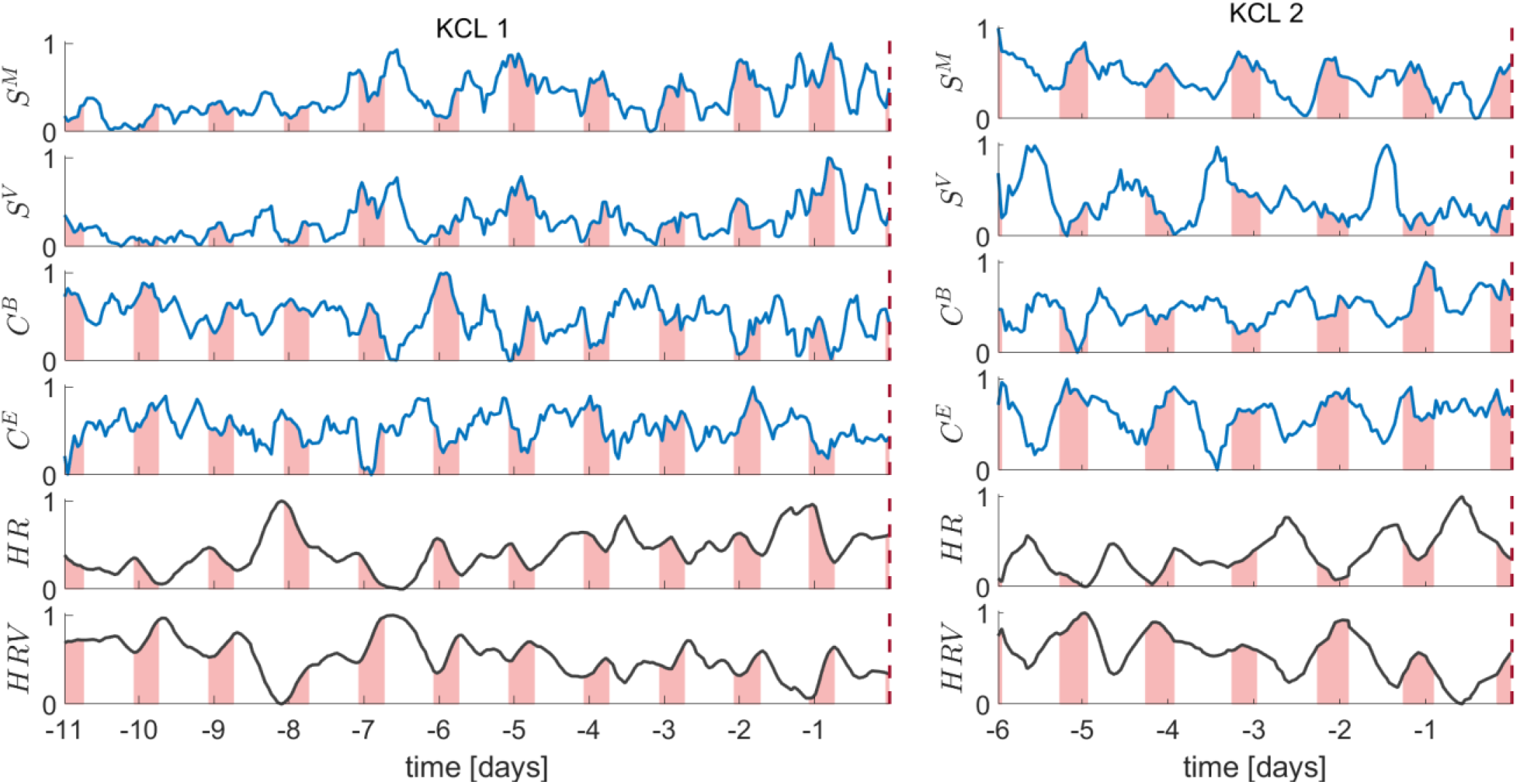
Temporal evolution of the EEG metrics in the alpha band (mean strength *S*^*M*^; variance of strength *S*^*V*^; average betweenness centrality *C*^*B*^; average eigenvector centrality *C*^*E*^) and ECG metrics (heart rate HR; heart rate variability HRV) for the KCL 1 and KCL 2 participants. Vertical dashed red line indicates the timestamp of the seizure. Shaded areas denote the 0am-8am time interval.

### 3.2. Periodicity of the EEG and ECG metrics

Figure 2 illustrates that the EEG and ECG metrics show a strong periodic behavior. To further investigate the contribution of different timescales in this apparent periodic behavior we computed the power spectral densities (Lomb-Scargle periodogram) of all metrics in all frequency bands. Figure 3 depicts the normalized power spectral densities of all metrics in the alpha frequency band for all study participants. We observe that the periodograms of the EEG metrics and particularly the variance of strength (*S*^*V*^) have in almost all participants a dominant peak around 24 hours. In addition, there is periodicity in the subharmonics around 12 and 8 hours. The ECG metrics show strong periodicity around 24 hours across all patients. Interestingly, similar results hold for the other frequency bands i.e., delta, theta, beta (Figures S1-S3). This suggests that the temporal evolution of EEG and ECG metrics is highly influenced by processes that occur on various timescales with strong contributions of daily rhythms.

**Figure 3.**
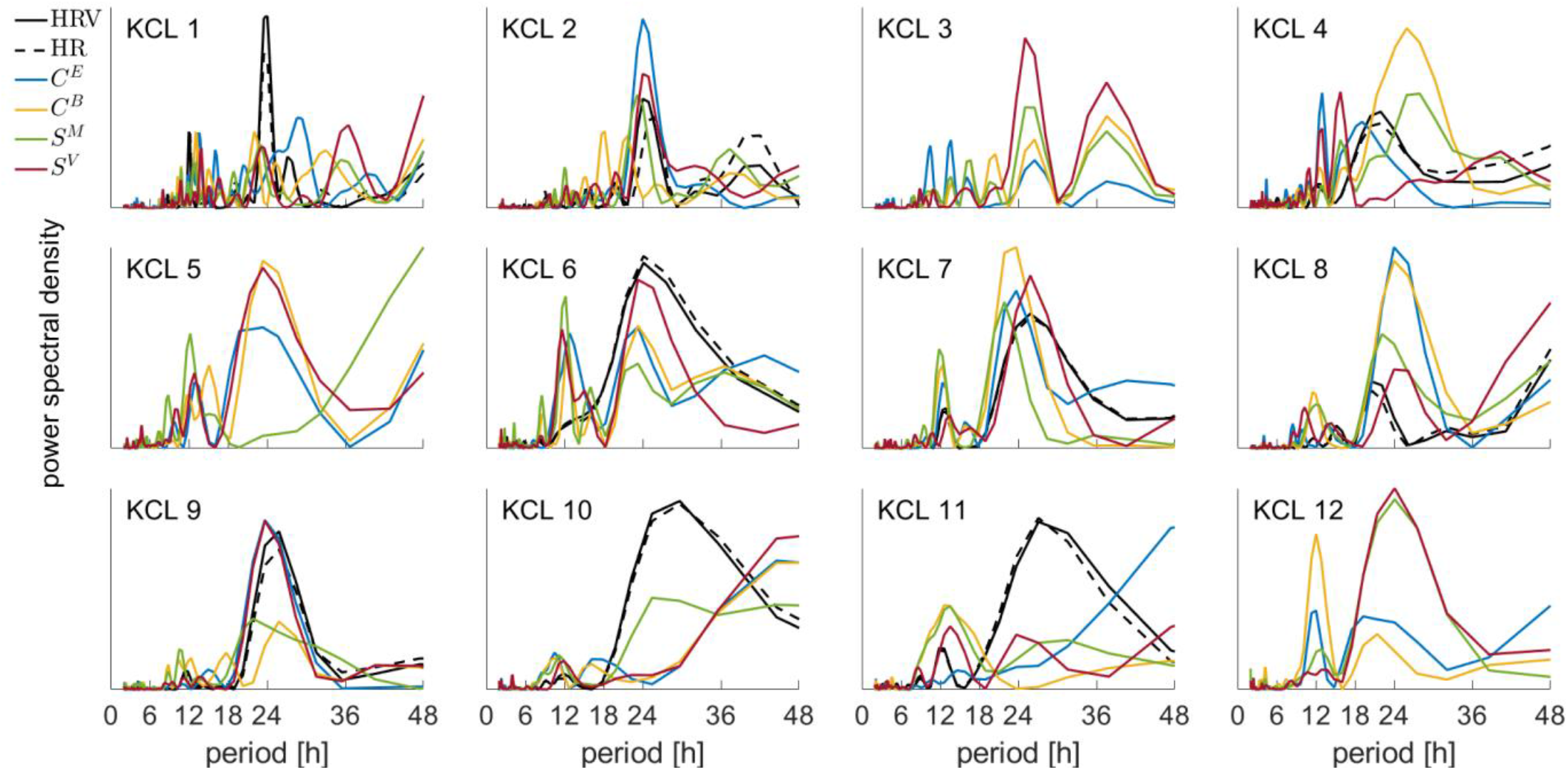
Periodograms of all the EEG and ECG metrics in the alpha frequency band. HRV: hear rate variability, HR: hear rate, *C*^*E*^: eigenvector centrality, *C*^*B*^: betweenness centrality, *S*^*M*^:mean strength, *S*^*V*^:variance of strength. Note that participants KCL 3, KCL 5 and KCL 12 do not have ECG metrics due to the absence of available ECG recordings (see Table 1).

### 3.3. Daily distributions of the EEG and ECG metrics

Having observed strong contributions of the daily rhythms in the temporal evolution of the EEG and ECG metrics (Figure 3) we sought to explore the temporal evolution of the daily distributions of the EEG and ECG metrics. In particular we investigated whether the distribution of the EEG/ECG metrics in the day prior to seizure occurrence is larger or smaller compared to the daily distributions of the corresponding metrics of all previous days. Hence, if a participant had five seizure-free days prior to seizure occurrence, i.e., {*d*_−1_, *d*_−2_, *d*_−3_, *d*_−4_, *d*_−5_} we obtained for each metric five daily distributions and applied four one-sided Wilcoxon rank sum tests (corrected for multiple comparisons, see Section 2.6) between the metrics of the day before the seizure *d*_−1_ and the metrics of each previous day i.e., {*d*_−2_, *d*_−3_, *d*_−4_, *d*_−5_}. We performed this analysis for all EEG/ECG metrics in all frequency bands.

Figure 4 illustrates the daily distributions of the variance of strength *S*^*V*^ in the alpha frequency band for all patients. We found that in 7 out of 12 participants the *S*^*V*^ distribution of the day before the seizure was either statistically significant larger (KCL 1, KCL 4, KCL 10, KCL 11) or smaller (KCL 5, KCL 8, KCL 12) compared to the *S*^*V*^distributions of all previous days. Interestingly, in 2 out of 11 patients (KCL 2, KCL 9) the *S*^*V*^ distributions of the two last days before the seizure occurrence were similar, but statistically significantly smaller to all previous days. In addition, in 3 out of 12 participants the daily distributions of the *S*^*V*^ values in the day before the seizure occurrence did not differ from the previous days. All p-values were corrected for multiple comparisons and reported in the Table S1.

**Figure 4.**
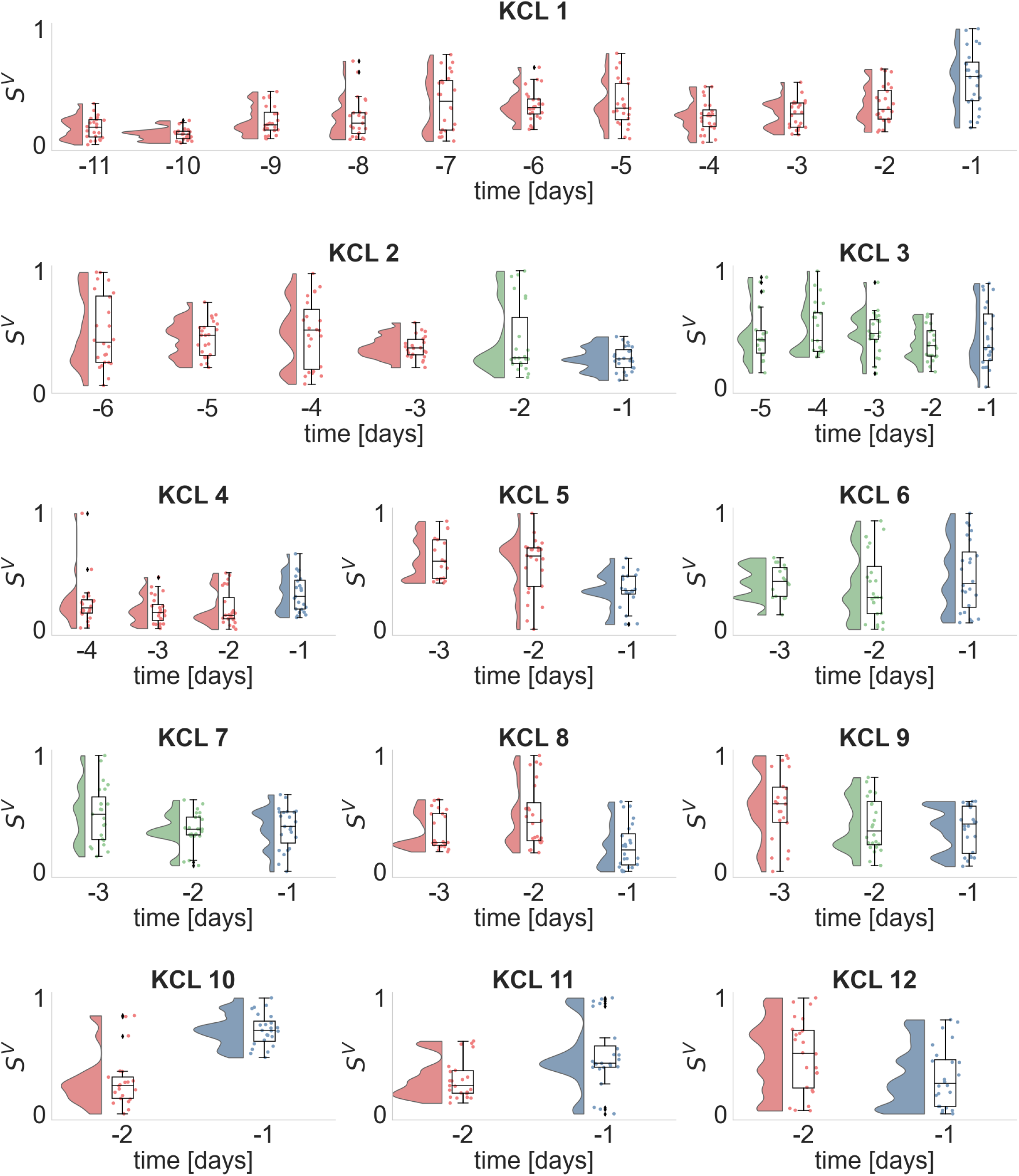
Daily distributions of the variance of strength *S*^*V*^ (alpha frequency band) for each seizure free day prior to seizure occurrence. Dots illustrate the *S*^*V*^ values obtained from each analyzed EEG segment, whilst histograms and boxplots depict their distribution. Horizontal lines in the boxplots indicate the median. The day before the seizure occurrence, i.e., *d*_−1_ is denoted with blue. Days whose distributions are statistically significantly different (one-sided Wilcoxon rank sum test) from the distribution of day *d*_−1_ are illustrated in red, otherwise are depicted in green.

We performed this analysis for the remaining frequency bands i.e. delta (Figure S4), theta (Figure S5) and beta (Figure S6), and found that the *S*^*V*^ distribution of the day prior to seizure occurrence was not consistently larger or smaller compared to the distributions of the previous days. Similar findings were also found for the mean strength (Figures S7-S10), average betweenness centrality (Figure S11-S14) and eigenvector centrality (Figure S15-S18) across all frequency bands. Therefore, the only informative EEG metric that showed significant changes in the EEG metrics daily distribution prior to seizure occurrence was the variance of strength in the alpha frequency band. To also ensure that these findings were not due to chance, we performed a bootstrap method by randomly shuffling 100 times the *S*^*V*^ values across different days (i.e., destroying the effect of the day whilst keeping the same number of points in each day). From this approach we excluded participants whose distributions were similar across days, i.e., KCL 3, KCL6 and KCL7. All p-values<0.05 and hence results were not attributed to chance.

Subsequently, we performed the same analysis for the ECG metrics. Figure 5 illustrates the daily distributions of the heart rate HR across the patients for whom there were available ECG recordings (Table 1). In 4 out of 9 participants the distribution of the day before the seizure was statistically significant larger (KCL 2, KCL7, KCL 9) or smaller (KCL 8) compared to the HR distributions of all previous days. In one out of 9 participants (KCL 4) the HR distributions of the two last days before the seizure occurrence were similar, but statistically significantly larger from the HR distribution of their previous day. For the remaining four participants either the daily HR distributions were similar across days (KCL 6, KCL 10, KCL 11) or there was a fluctuation in the HR values across days (KCL 1). The corresponding heart rate variability (HRV) distributions were qualitatively similar however with opposite direction from the HR distributions (Figure S19).

**Figure 5.**
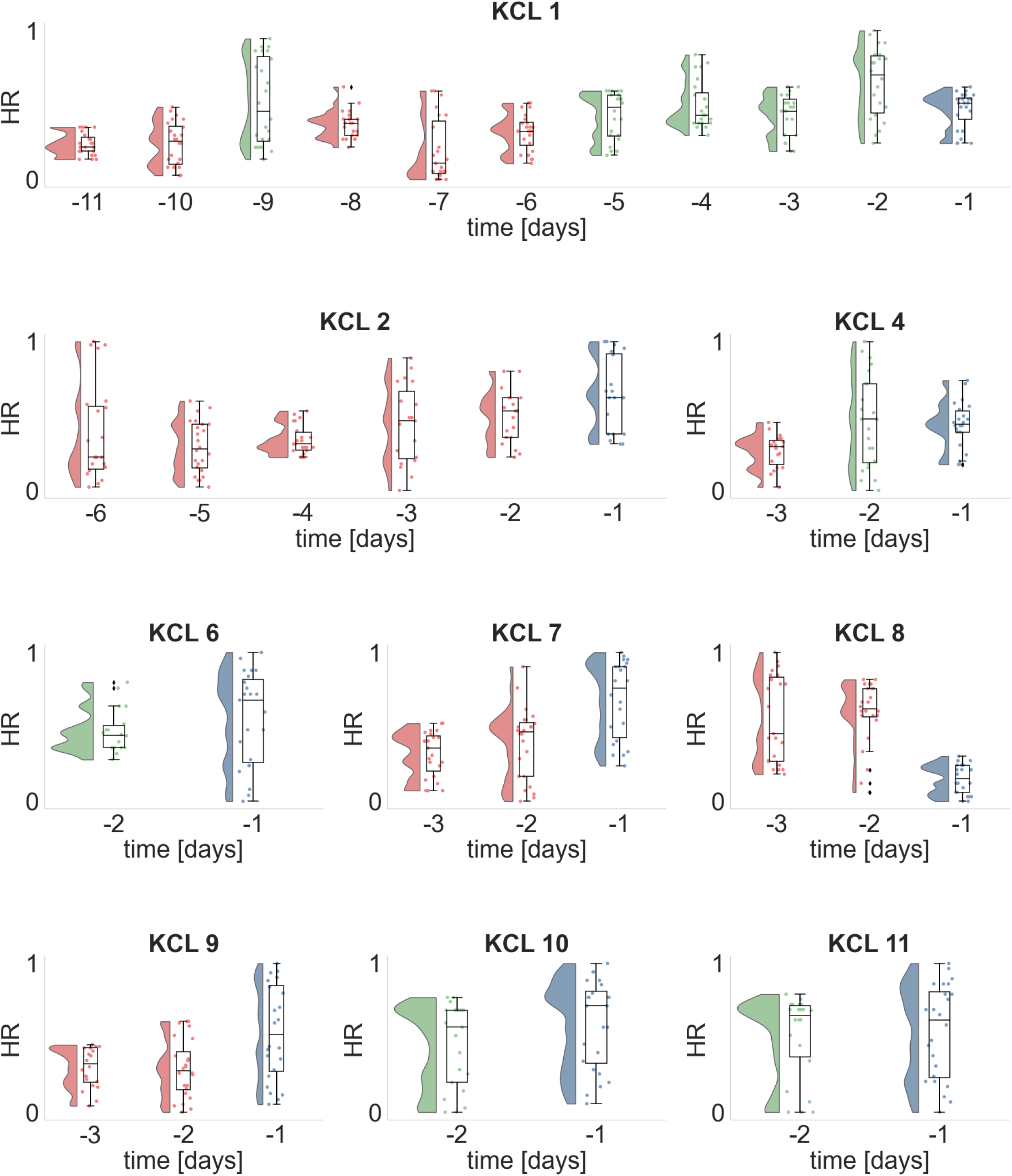
Daily distributions of the heart rate HR for each seizure free day prior to seizure occurrence. Dots illustrate the HR values obtained from each analyzed ECG segment, whilst histograms and boxplots depict their distribution. Horizontal lines in the boxplots indicate the median. The day before the seizure occurrence, i.e., *d*_−1_ is denoted with blue. Days whose distributions are statistically significantly different (one-sided Wilcoxon rank sum test) from the distribution of day *d*_−1_ are illustrated in red, otherwise are depicted in green.

### 3.4. Lateralization of the Seizure Focus

Having investigated the temporal evolution of the daily distributions of the EEG metrics prior to seizures and found that the alpha frequency band is the most informative, we sought to explore whether we can use the EEG metrics in the alpha band for further analysis. We investigated whether the analyzed data could be informative to lateralize the hemisphere of the seizure focus. Five study participants had a clear seizure focus (Table 1; KCL 1, KCL 8 had a seizure focus on the right hemisphere; KCL 4, KCL 5, KCL 6 had the seizure focus on the left hemisphere). Figure 6 depicts the distributions of the average eigenvector centrality of each hemisphere across all analyzed patients. We found that in four out of five patients the hemisphere that contained the seizure focus had statistically significantly higher eigenvector centrality compared to the other hemisphere. We performed this analysis for the other EEG metrics i.e., mean strength, variance of strength, betweenness centrality, however, in mean strength and variance of strength there were at least 3 out of 5 patients for which there was not a statistically significant difference between the distributions of the two hemispheres (Figure S20 and S21). In addition, in the average between centrality in 2 out of 5 participants there were statistically significantly differences between the two hemispheres and in one participant the difference was in another direction (Figure S22)

**Figure 6.**
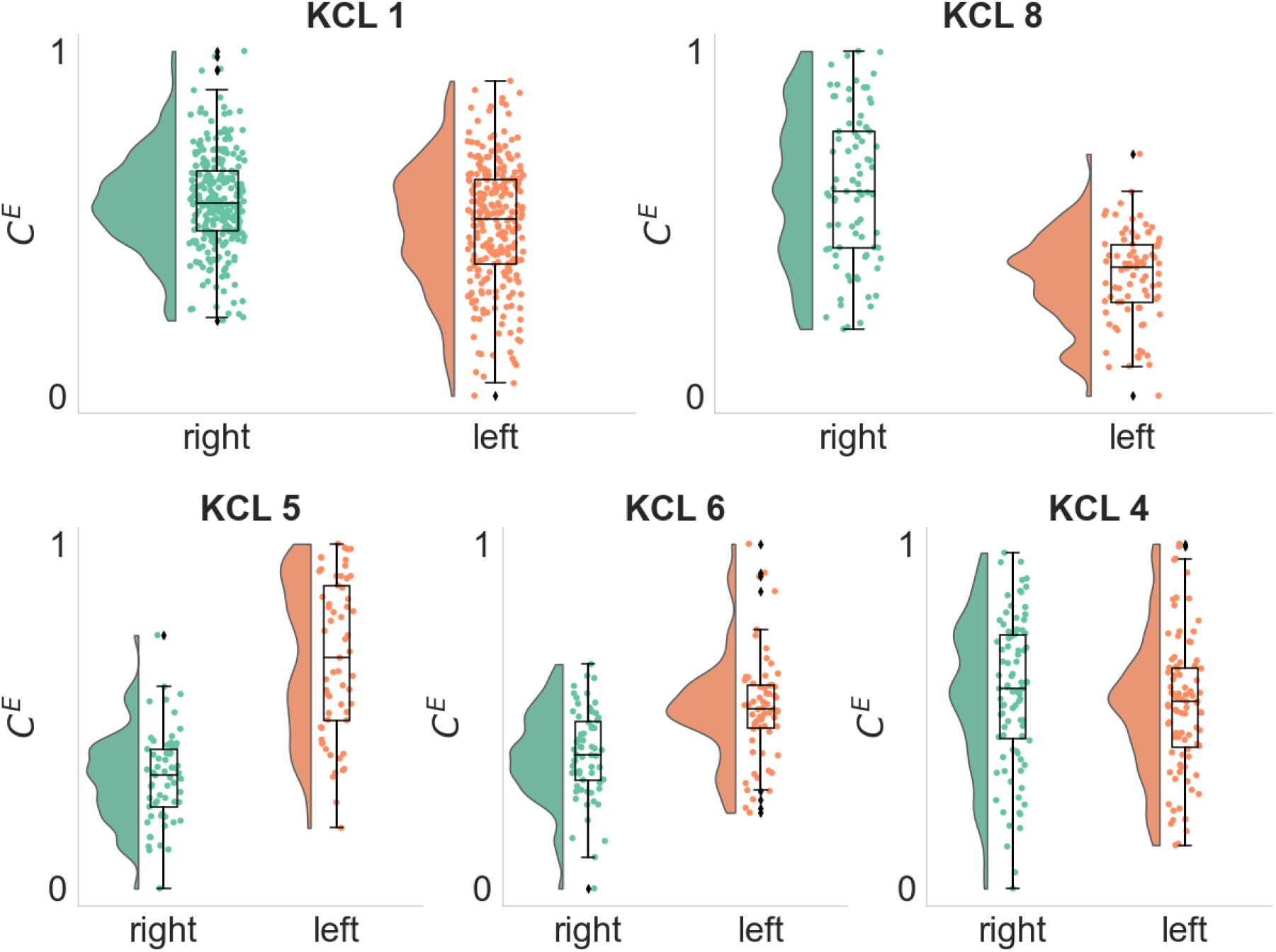
Distributions of the Average Eigenvector Centrality *C*^*E*^ of each hemisphere in the alpha frequency band. Each dot depicts the *C*^*E*^ value of the right (green) or left (orange) hemisphere as computed from a single EEG segment. KCL 1 and KCL 2 participants had the seizure focus on the right hemisphere (p-values 3.13 × 10^−6^ and 3.54 × 10^−13^ respectively, one-sided Wilcoxon rank-sum test), whilst KCL 5, KCL 6 and KCL 4 had the seizure focus on the left hemisphere (p-values 1.75 × 10^−14^, 3.63 × 10^−7^ and 0.9 respectively, one-sided Wilcoxon rank-sum test).

## 4. Discussion

In this study we examined the multiday temporal evolution of brain network metrics prior to seizure occurrence. Using samples of scalp EEG recordings from every single hour of each day we inferred functional brain networks in four frequency bands across multiple days. We quantified the topological properties of the brain networks using metrics from graph theory, i.e., mean strength, variance of strength, average betweenness centrality and average eigenvector centrality. We found that the brain network metrics fluctuate over time (Figure 2) and exhibit periodic-like behaviour with prominent circadian features (Figures 3, S1-S3). Considering the contribution of daily rhythms in the temporal evolution of the brain network metrics, we investigated whether the metrics in the day prior to seizure were statistically significantly larger or smaller compared to all previous days. We found that in 7 out of 12 patients the variance of strength in the alpha band on the day prior to seizure was significantly different to all previous days; and in 2 out of 12 patients the two last days prior to the seizure had similar distributions but were significantly different to all previous days (Figure 4). Having observed that the alpha frequency band is the most informative band in the temporal evolution of brain network metrics, we sought to explore whether the graph metrics can be used to lateralize the seizure focus in the alpha band. We found that in 4 out 5 patients who had a clear seizure focus the eigenvector centrality was able to lateralize the hemisphere of the seizure focus (Figure 6).

Tools from network neuroscience have been widely used to characterize topological properties of functional networks inferred from EEG recordings [Bassett_2018]. In the context of epilepsy, Khambhati et al. [Khambhati_2021] showed that long-term frequency-dependent reorganization of interictal functional networks reflects seizure proneness. In addition, Chowdhury at al. 2014 [Chowdhury_2014] showed that mean degree (equivalent to strength) in the low alpha band is able to distringuish PWE from controls whilst Pegg et al. [Pegg_2020] showed that mean degree can distinguish people with well-controlled and drug-resistant focal epilepsy. Furthermore, variance of strength has been reported to be higher in PWE and their first-degree relatives compared to controls [Chowdhury_2014]. Eigenvector centrality is closely associated to computational metrics that quantify the tendency of a brain network to generate seizures and it is able to identify brain regions responsible for seizure generation [Lopes_2017, Laiou_2019]. Moreover, betweenness centrality has been reported as a useful metric to identify brain regions that are neighboring to the seizure onset zone [Geier_2015]. Hence, we used in this study four network metrics i.e. mean strength (quantifies network connectedness), variance of strength (variation in the node strength distribution), average eigenvector centrality (quantifies presence of network hubs) and average betweenness centrality (quantifies presence of nodes that act like network bridges), to capture different topological properties of the functional networks.

We found that the brain network metrics fluctuate over time and show periodic behaviour with a peak period around 24 hours (Figures 2, 3, S1-S3). These findings are in line with previous studies that investigated the temporal evolution of brain network metrics using intracranial EEG recordings. Kuhnert et al. [Kuhnert_2010] and Geier et al. [Geier_2017] inferred functional brain networks and used strength, betweeness centrality, clustering coefficient, and shortest path length to characterize the topological properties of the networks. They reported temporal fluctuations of the metrics over time to a greater or lesser extent as well as the strong contribution of daily rhythms in this temporal variation. However, those studies did not perform their analysis on specific frequency bands but applied a broadband filtering.

When we examined the temporal evolution of the daily distributions of brain network metrics, we asked whether the metrics distribution in the day prior to seizure takes smaller or larger values compared to all previous days (Figure 3, S4-S19), and found that the most informative metric was the variance of strength in the alpha frequency band (Figure 3). In 7 out of 12 participants the distribution of the variance of strength in the day prior to seizure was consistently larger (4 participants) or smaller (3 participants) compared to all previous days. In 2 out of 12 patients the variance of strength distributions of the two last days prior to seizure were similar but statistically significantly smaller to all previous days. The variance of strength quantifies the variability of the strength values across the network nodes. High variance of strength means that the network has nodes whose strength is much higher than the average strength of the network. This might indicate a more regular network topology with nodes that act like hubs. On the other hand, low variance of strength means that the strength values of across nodes are similar. Lower strength variability could be linked to increased synchronization (i.e., all nodes tend to connect together tightly and have similar large strength values) or decrease in syncrhonization (i.e., the connections between the nodes become more loose and all nodes have similar small strength values). It is suspected that seizures are generated due to the imbalance between inhibition and excitation [Jiruska_2013]. Previous studies that used intracranial EEG recordings and studied changes in phase synchronization and network topology metrics before, during and after seizure in a high temporal resolution (from seconds and minutes to hours) reported both decrease and increase in synchronization prior to seizure occurrence, however, a synchronization decrease is the most prevalent [Mormann_2005, Le Van Quyen_2005, Jiruska_2013]. In addition, Schindler at al. [Schindler_2008] reported that the network topology changes from a more random structure to more regular in seizures and returns back to randomness towards seizure termination.

In patients with a clear seizure focus we also found that average eigenvector centrality was higher in the hemisphere of the seizure focus (Figure 6). Eigenvector centrality is a measure of node importance and in the context of epilepsy surgery it has been shown to serve a useful marker for the identification of network hubs and brain regions responsible for seizure generation [Lopes_2017, Laiou_2019]. In addition, Coito et al. 2015 [Coito_2015] studied directed connectivity in patients with left and right temporal lobe epilepsy and found different patterns of time-varying connectivity between the two groups as well as concordance of the highest outlflow region with the epileptogenic zone.

Considering that the EEG acquisition occurred simultaneously with an ECG acquisition we also investigated the multiday temporal evolution of heart rate and heart rate variability metrics. We quantified the heart rate variability using the root mean square of the successive difference of R-peaks (RMSSD). Previous studies have shown that RMSSD correlates with ECG frequency metrics, and it is a realible metric even in ultra short-term ECG recordings [Shaffer_2017, Shaffer_2020]. We found that the HR and HRV have a strong periodicity around 24 hours (Figure 3). This is in line with recent work from Karoly at al. [Karoly_2021] and Gregg at al. 2022 [Gregg_2022] that used wearable devices to investigate heart rate cycles in people with epilepsy and found the presence of circadian and multiday cycles. In addition, we found that in 4 out of 9 participants the HR distribution of the day before the seizure was statistically significant larger (3 participants) or smaller (1 participant) compared to the distributions of all previous days. In one out of 9 participants (KCL 4) the HR distributions of the two last days before the seizure occurrence were similar, but statistically significantly larger from the HR distribution of their previous day. The corresponding heart rate variability (HRV) distributions were qualitatively similar with the HR distributions however with opposite trend (Figure S19).

Our findings should be interpreted by taking into account the study limitations. First, the study participants were under combination therapy with two or more antiepileptic drugs (AEDs) and during their stay in the EMU there was an ongoing change in the AEDs in a patient specific manner. In addition, the analysed cohort was small and hence further studies are needed to extend and validate our findings.

In conclusion the findings of this study suggest that multiday brain network metrics computed from EEG recordings could potentially be used to characterize brain network changes that occur prior to seizures. In addition, they show promise as potential biomarkers for the estimation of seizure risk and their incorporation in the study pipelines for a long-term wearable EEG mobile monitoring for epilepsy forecasting and management.

## Data Availability

All data produced in the present study are available upon reasonable request to the RADAR-consortium

## Supplementary Materials

The Supplementary material contains Figures (Figure S1-S22) and a table (Table S1)

## Author Contributions

“Conceptualization, P.L., A.B., M.P.R; methodology, P.L., M.D.; software, P.L.; validation, P.L., M.D.; formal analysis, P.L.; data curation, A.B., E.B., P.V., J.S.W.; writing—original draft preparation, P.L., A.B.; writing—review and editing, P.L, A.B., E.B., P.V., J.S.W., Z.R., Y.R., P.C., C.S., S.S., Y.Z., A.F., R.J.B.D., A.S.B., M.D., M.P.R; visualization, P.L., A.B.; supervision, M.P.R.; funding acquisition, M.P.R., A.S.B., R.J.B.D.

All authors have read and agreed to the published version of the manuscript.”

## Funding

The Remote Assessment of Disease and Relapse–Central Nervous System project has received funding from the Innovative Medicines Initiative 2 Joint Undertaking under grant agreement number 115902. This Joint Undertaking receives support from the European Union’s Horizon 2020 research and innovation programme and European Federation of Pharmaceutical Industries and Associations [31]. This communication reflects the views of the Remote Assessment of Disease and Relapse–Central Nervous System consortium and neither Innovative Medicines Initiative nor the European Union and European Federation of Pharmaceutical Industries and Associations are liable for any use that may be made of the information contained herein. The funding body have not been involved in the design of the study, the collection or analysis of data, or the interpretation of data. This paper represents independent research part funded by the National Institute for Health Research (NIHR) Maudsley Biomedical Research Centre at South London and Maudsley National Health Service (NHS) Foundation Trust and King’s College London. The views expressed are those of the authors and not necessarily those of the NHS, the NIHR, or the Department of Health and Social Care.

## Acknowledgments

The authors are grateful to Drs Kasper Claes, Valentina Ticcinelli, Nikolay Manyakov and Srinivasan Vairavan for useful discussions.

## Conflicts of Interest

The authors declare no conflict of interest. In addition, the funders had no role in the design of the study; in the collection, analyses, or interpretation of data; in the writing of the manuscript, or in the decision to publish the results.

## Notes

### Competing Interest Statement

The authors have declared no competing interest.

