## Supplementary material for "Temporal evolution of multiday, epileptic functional networks prior to seizure occurrence": Supplementary_material_MedRxiV.pdf

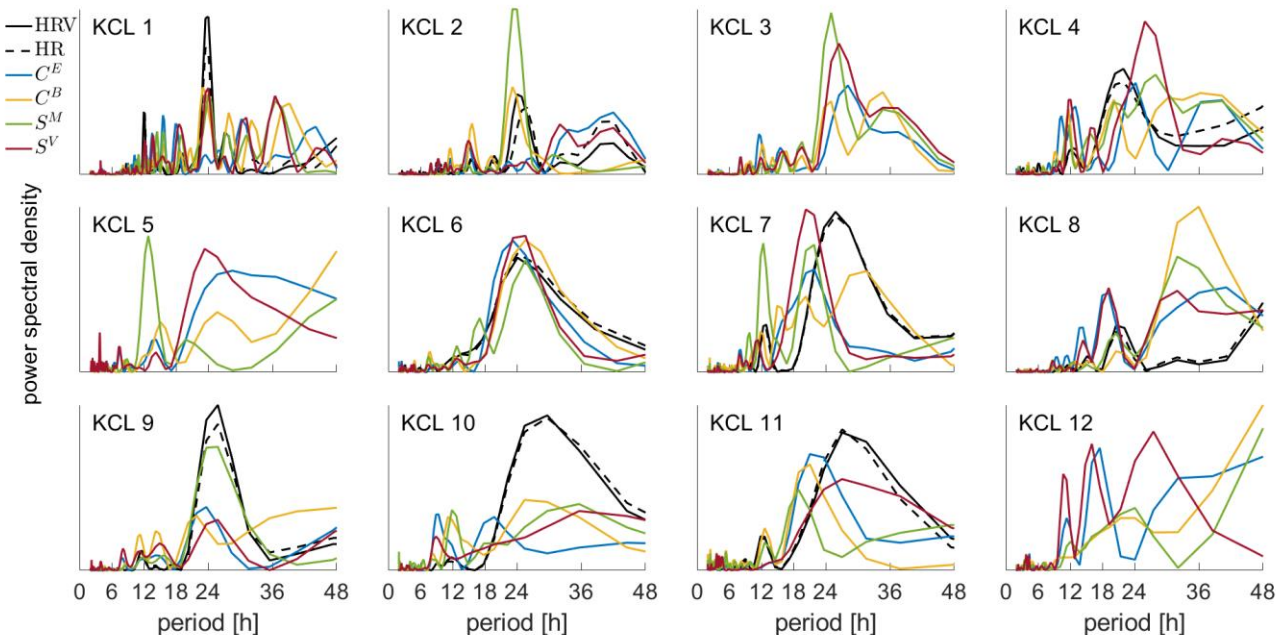


**Figure S1**. Periodograms of all the EEG and ECG metrics in the delta frequency band. HRV: heart rate variability, HR: heart rate, $C^E$: eigenvector centrality, $C^B$: betweenness centrality, $S^M$:mean strength, $S^V$:variance of strength. Note that participants KCL 3, KCL 5 and KCL 12 do not have ECG metrics due to the absence of available ECG recordings (see Table 1).

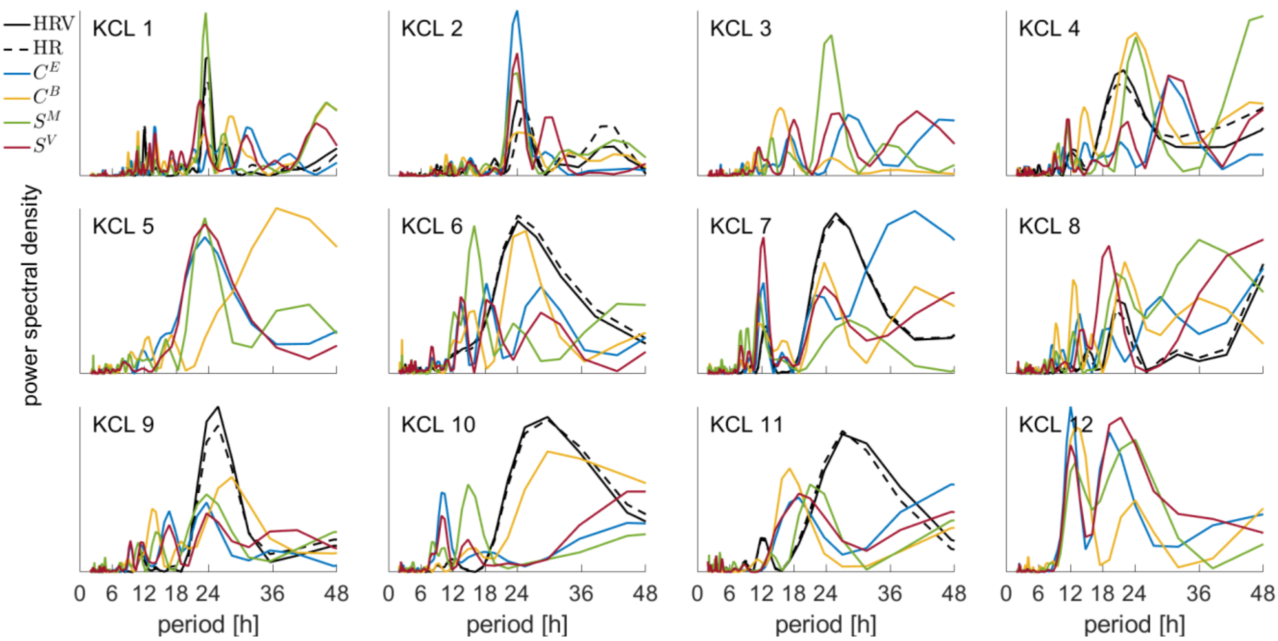


**Figure S2**. Same as Figure S1, but in the theta band.

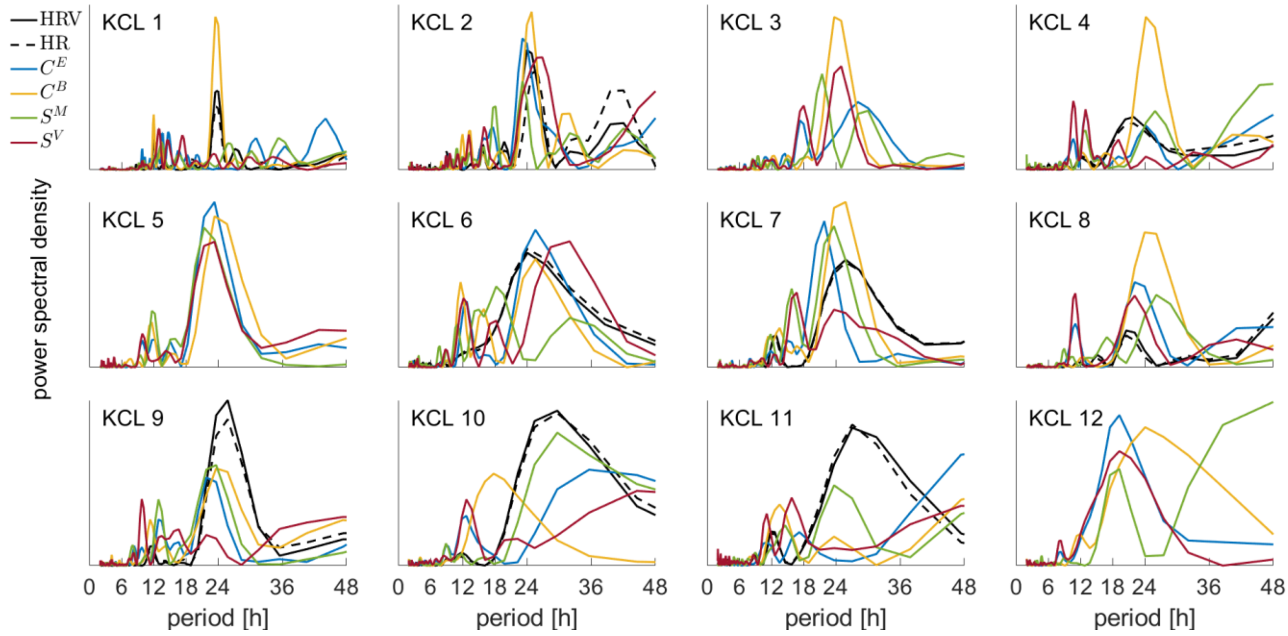


**Figure S3**. Same as Figure S1, but in the beta band.

**Table S1**. Table with the original p-values (one-sided Wilcoxon rank-sum test) between the variance of strength distribution of the day prior to seizure ($d_{-1}$) and the corresponding distribution of all previous days.

| | $d_{-11,-1}$ | $d_{-10,-1}$ | $d_{-9,-1}$ | $d_{-8,-1}$ | $d_{-7,-1}$ | $d_{-6,-1}$ | $d_{-5,-1}$ | $d_{-4,-1}$ | $d_{-3,-1}$ | $d_{-2,-1}$ |
|---|---|---|---|---|---|---|---|---|---|---|
| KCL 1 | **$9 \times 10^{-7}$** | **$3\text{x}10^{-8}$** | **$10^{-5}$** | **$11 \times 10^{-4}$** | **0.0054** | **0.0008** | **0.0020** | **$10^{-4}$** | **$8\text{x}10^{-5}$** | **0.0013** |
| KCL 2 | | | | | | **0.0065** | **0.0001** | **0.0049** | **0.0012** | 0.1365 |
| KCL 3 | | | | | | | 0.3191 | 0.1431 | 0.1335 | 0.6334 |
| KCL 4 | | | | | | | | **0.0296** | **0.0008** | **0.0011** |
| KCL 5 | | | | | | | | | **$7\text{x}10^{-5}$** | **$8\text{x}10^{-4}$** |
| KCL 6 | | | | | | | | | 0.6350 | 0.8861 |
| KCL 7 | | | | | | | | | 0.0620 | 0.7835 |
| KCL 8 | | | | | | | | | **0.0070** | **0.0002** |
| KCL 9 | | | | | | | | | **0.0090** | 0.3363 |
| KCL 10 | | | | | | | | | | $1.3\text{x}10^{-6}$ |
| KCL 11 | | | | | | | | | | **0.0016** |
| KCL 12 | | | | | | | | | | **0.0191** |

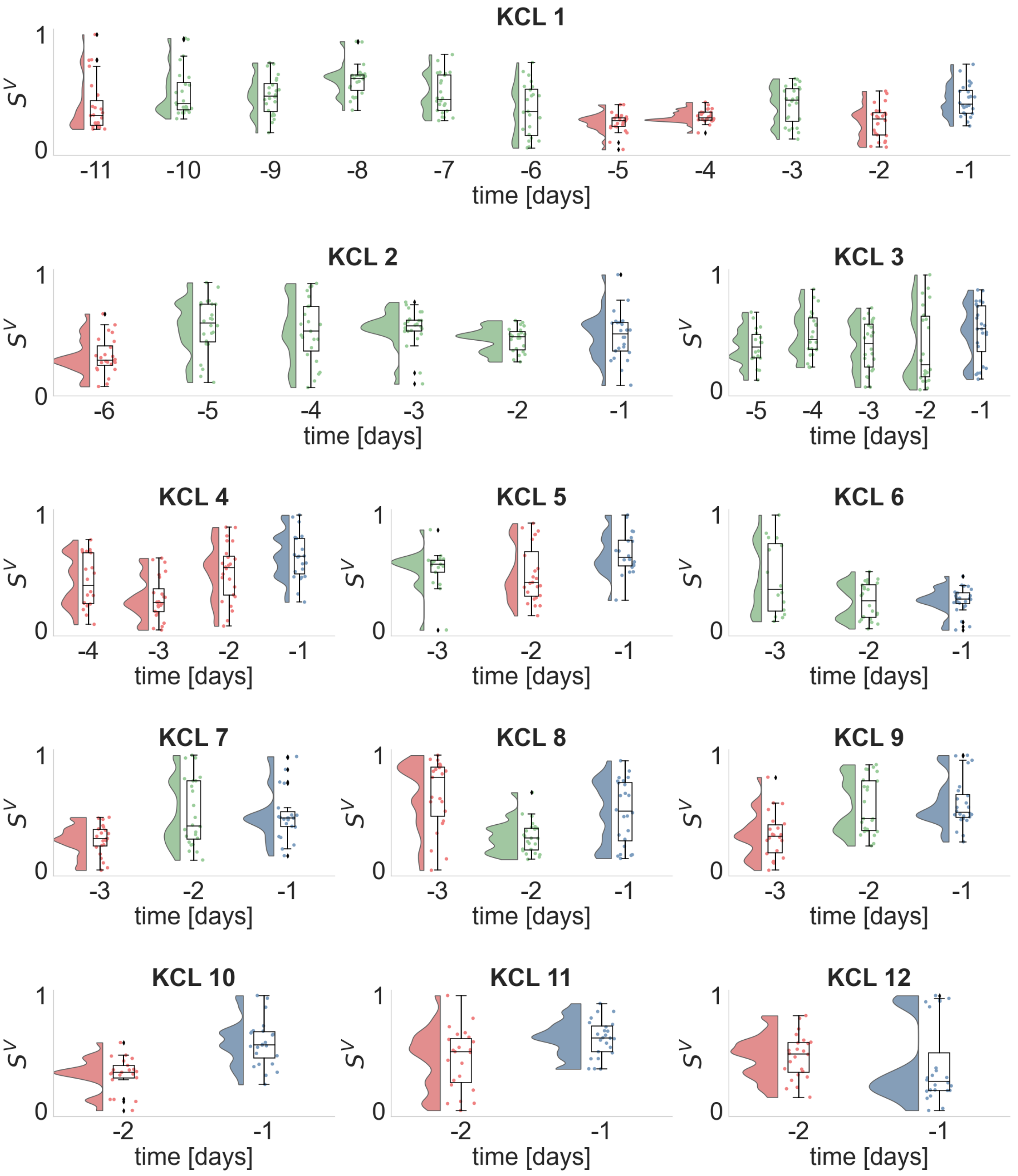


**Figure S4**. Daily distributions of the variance of strength $S^V$ (delta frequency band) for each seizure free day prior to seizure occurrence. Dots illustrate the $S^V$ values obtained from each analyzed EEG segment, whilst histograms and boxplots depict their distribution. Horizontal lines in the boxplots indicate the median. The day before the seizure occurrence, i.e., $d_{-1}$ is denoted with blue. Days whose distributions are statistically significantly different (one-sided Wilcoxon rank sum test) from the distribution of day $d_{-1}$ are illustrated in red, otherwise are depicted in green.

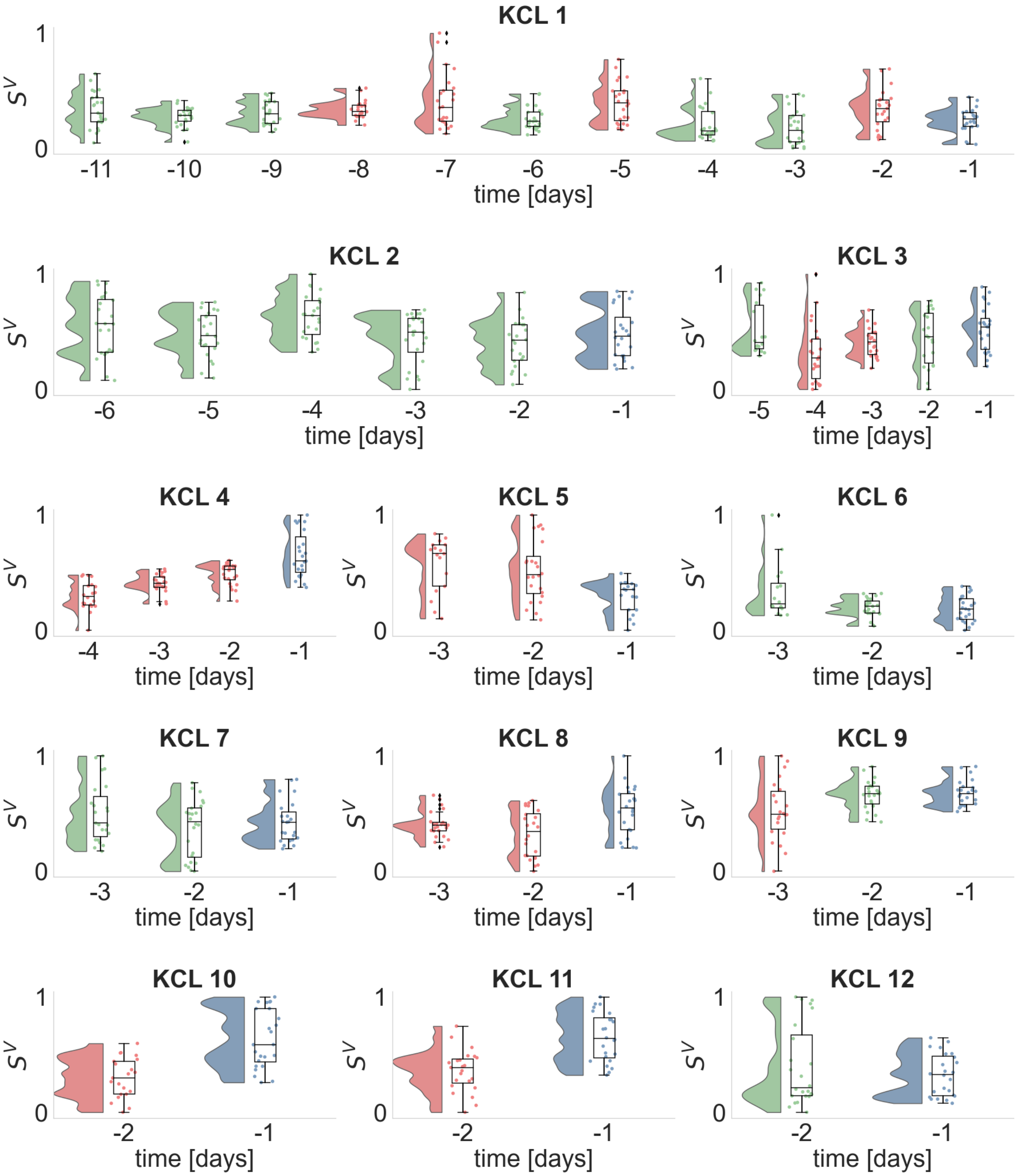


**Figure S5**. Same as Figure S4 for the theta band.

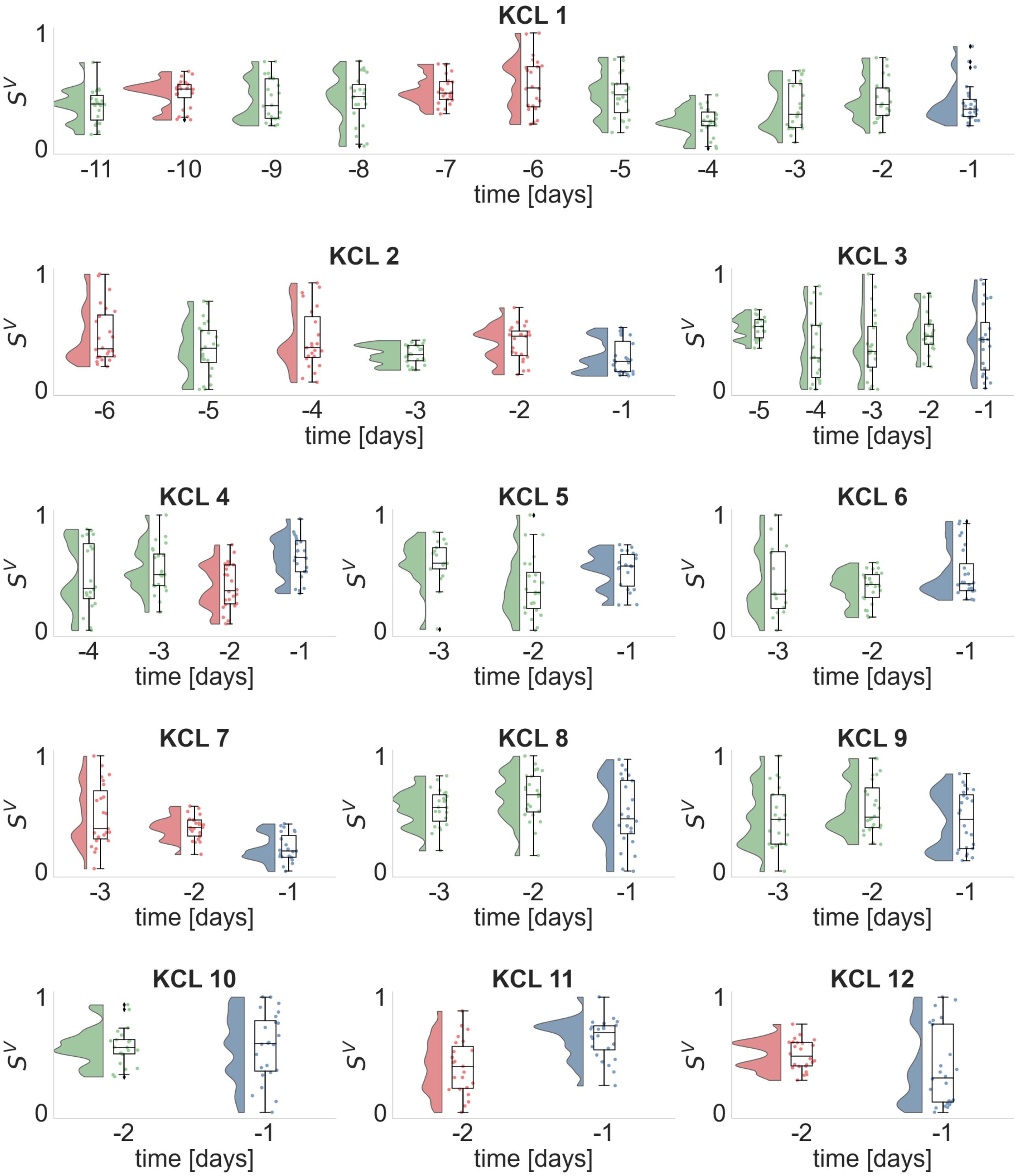


**Figure S6**. Same as Figure S4 for the beta band.

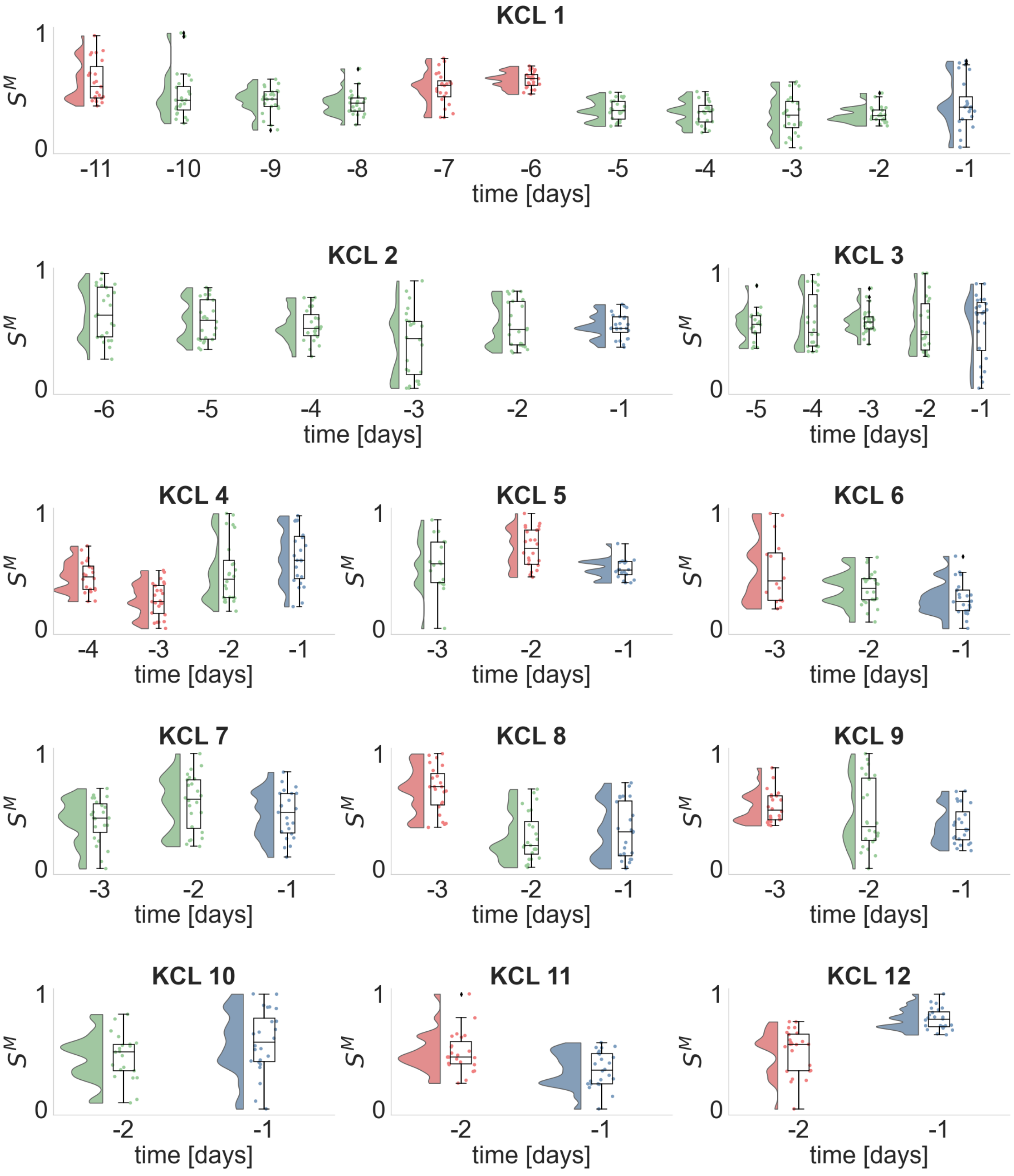


**Figure S7**. Same as Figure S4 for the mean strength and delta band.

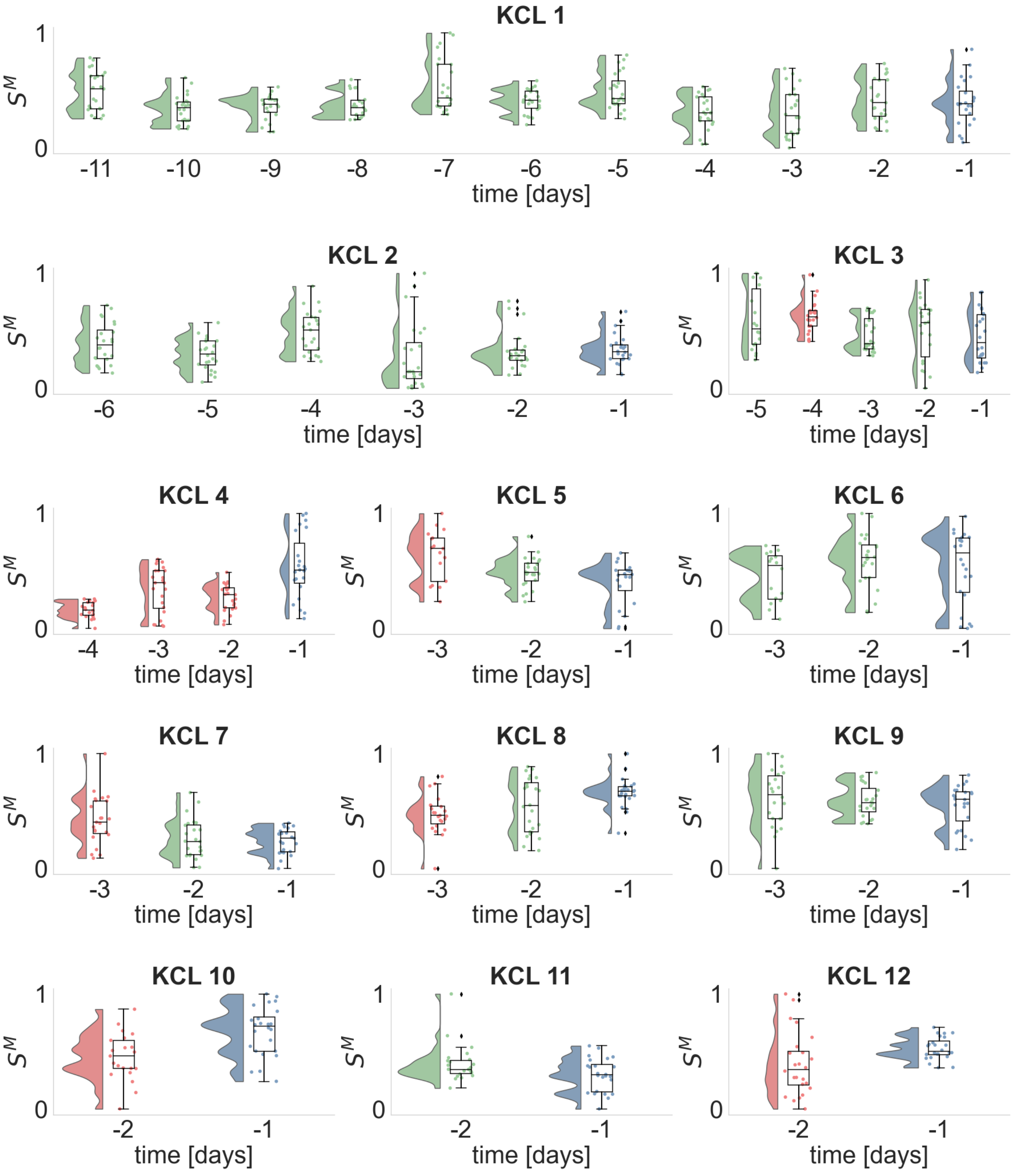


**Figure S8**. Same as Figure S4 for the mean strength and theta band.

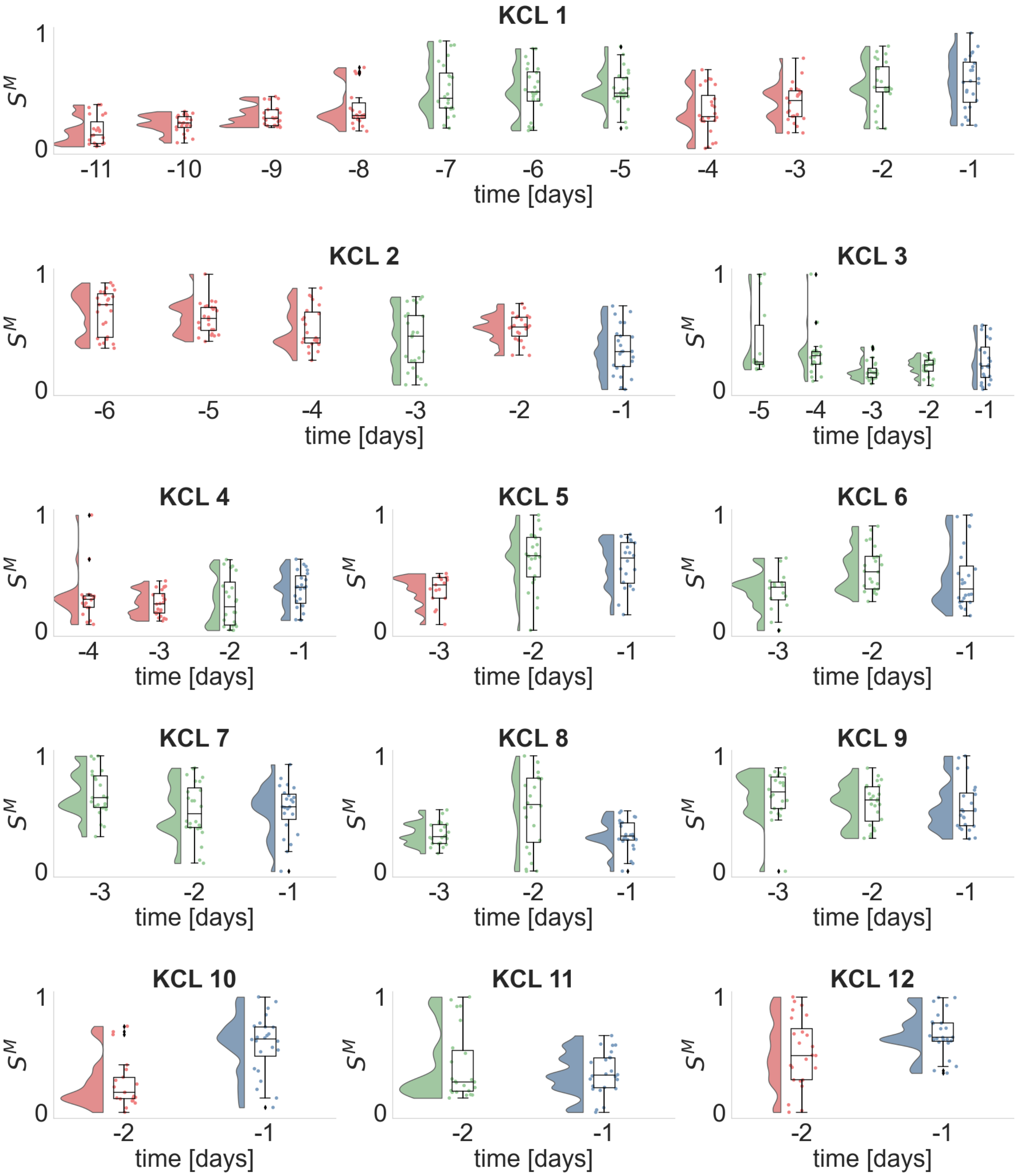


**Figure S9**. Same as Figure S4 for the mean strength and alpha band.

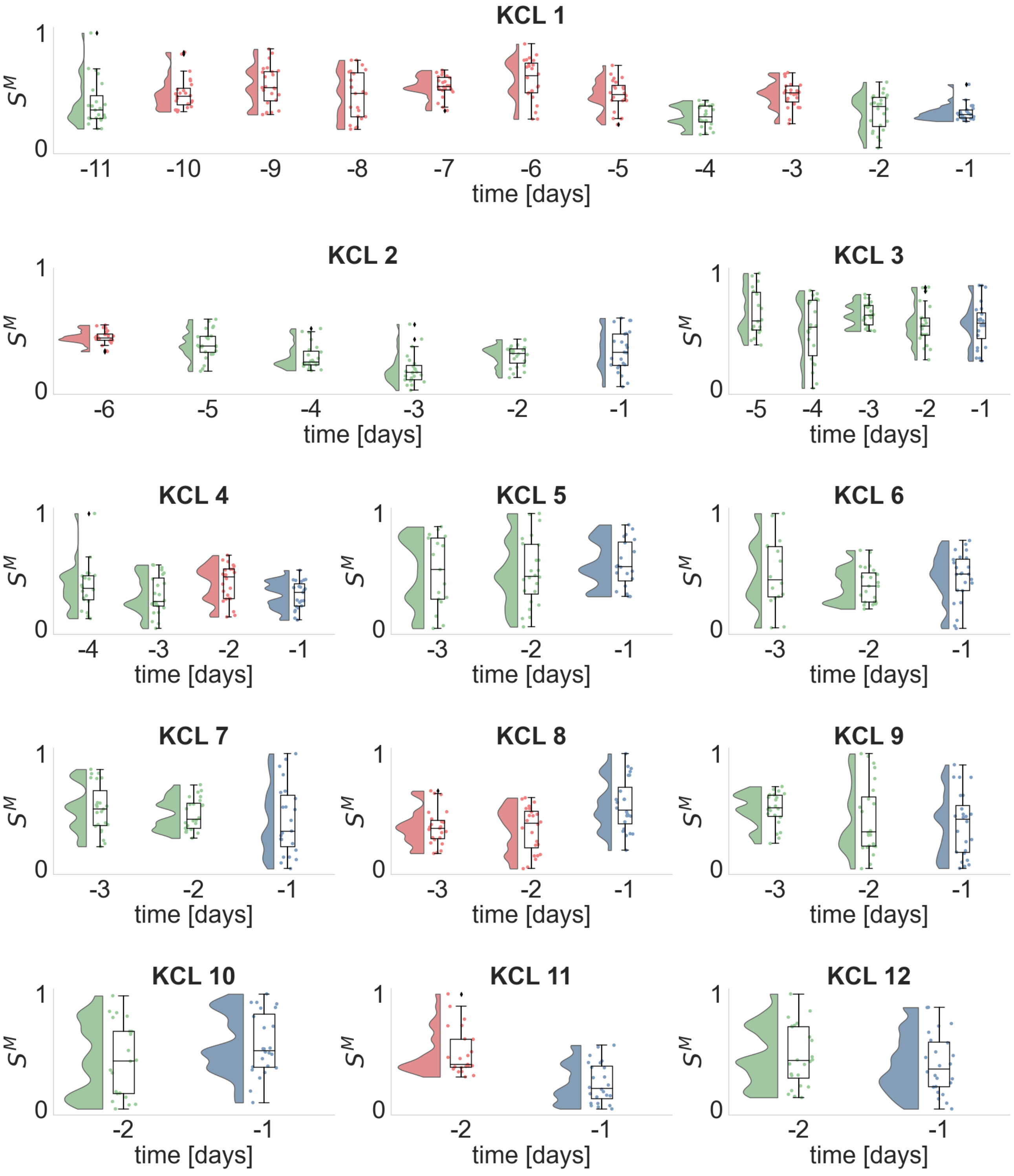


**Figure S10**. Same as Figure S4 for the mean strength and beta band.

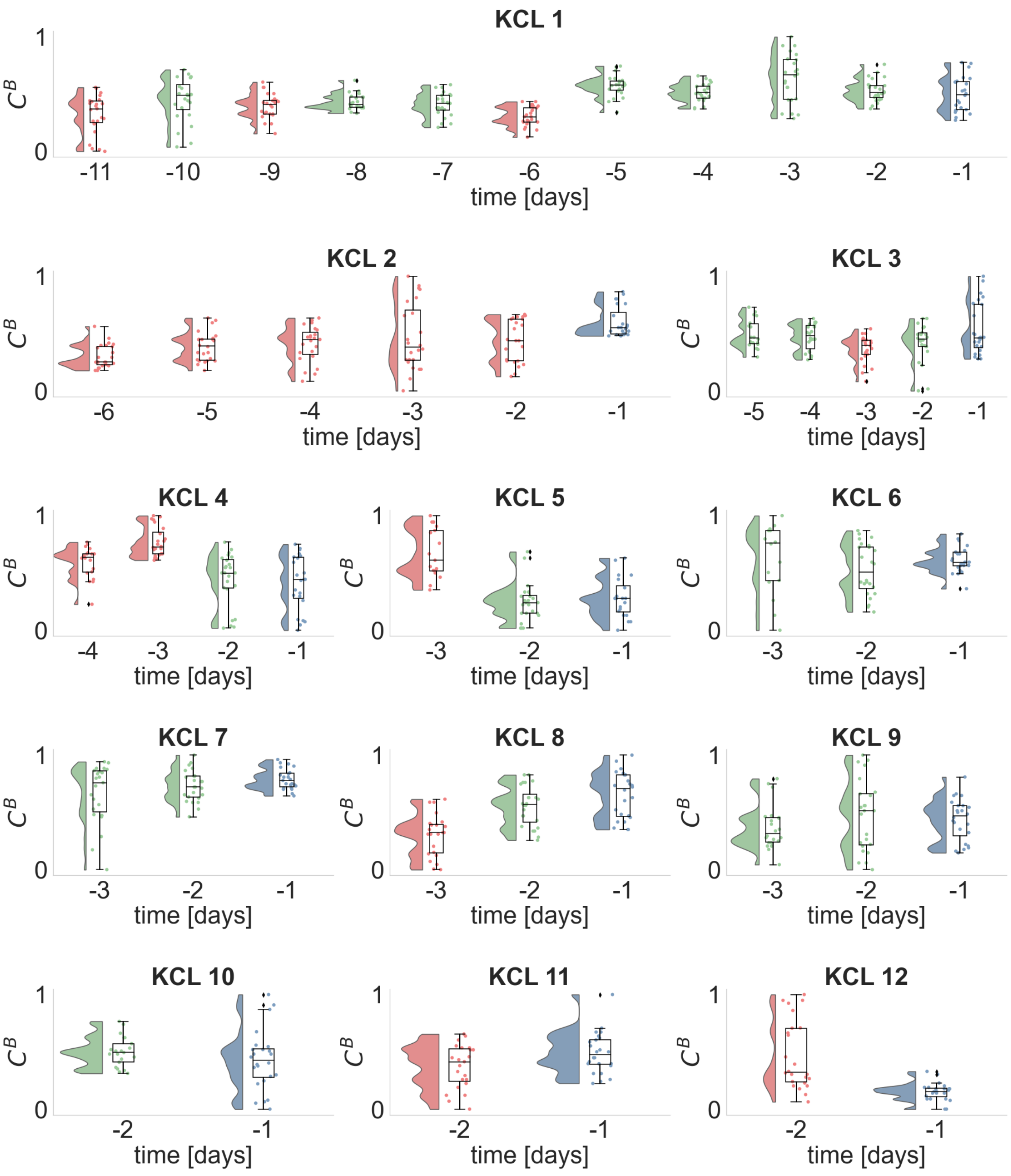


**Figure S11**. Same as Figure S4 for the average betweenness centrality and delta band.

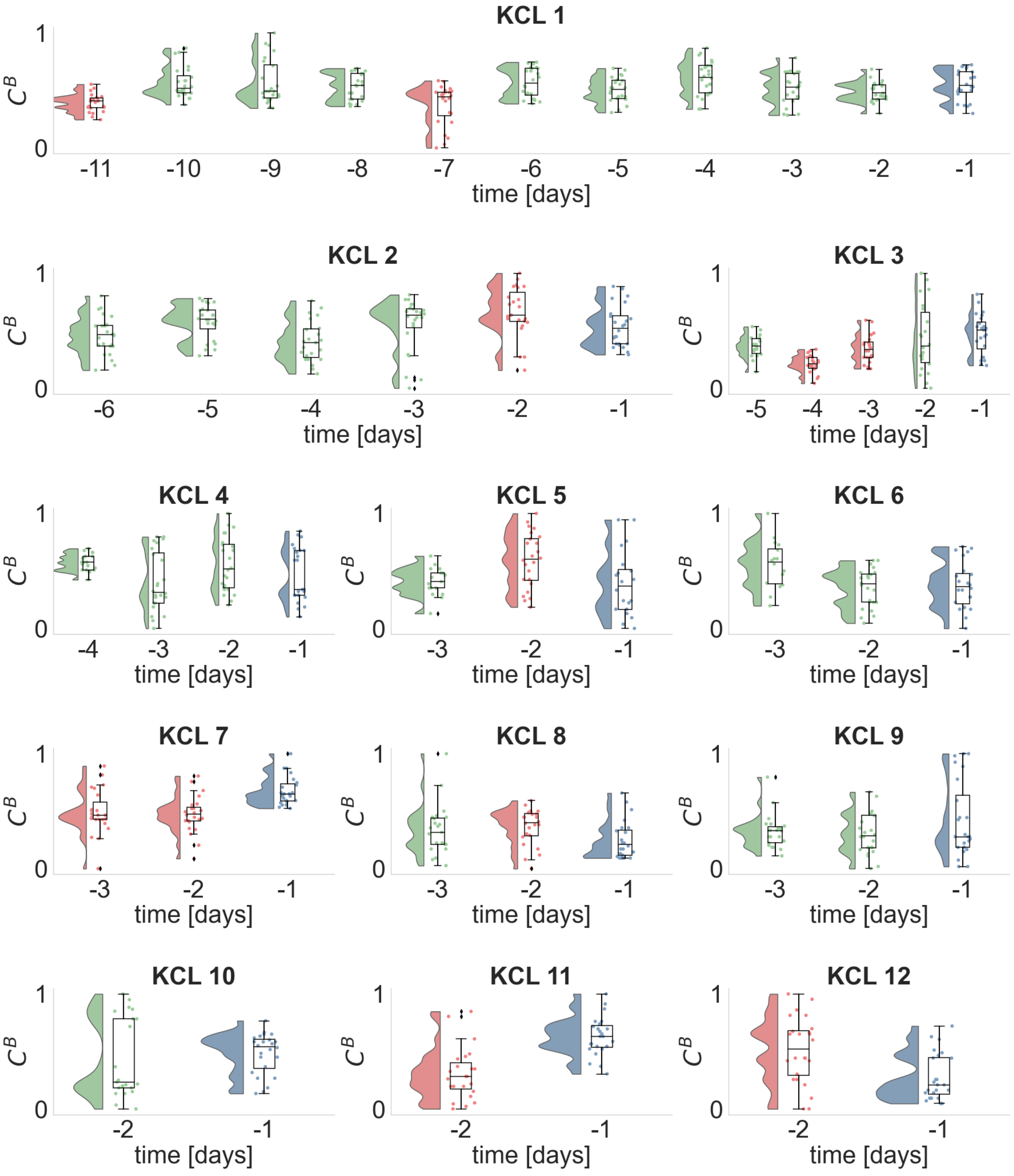


**Figure S12**. Same as Figure S4 for the average betweenness centrality and theta band.

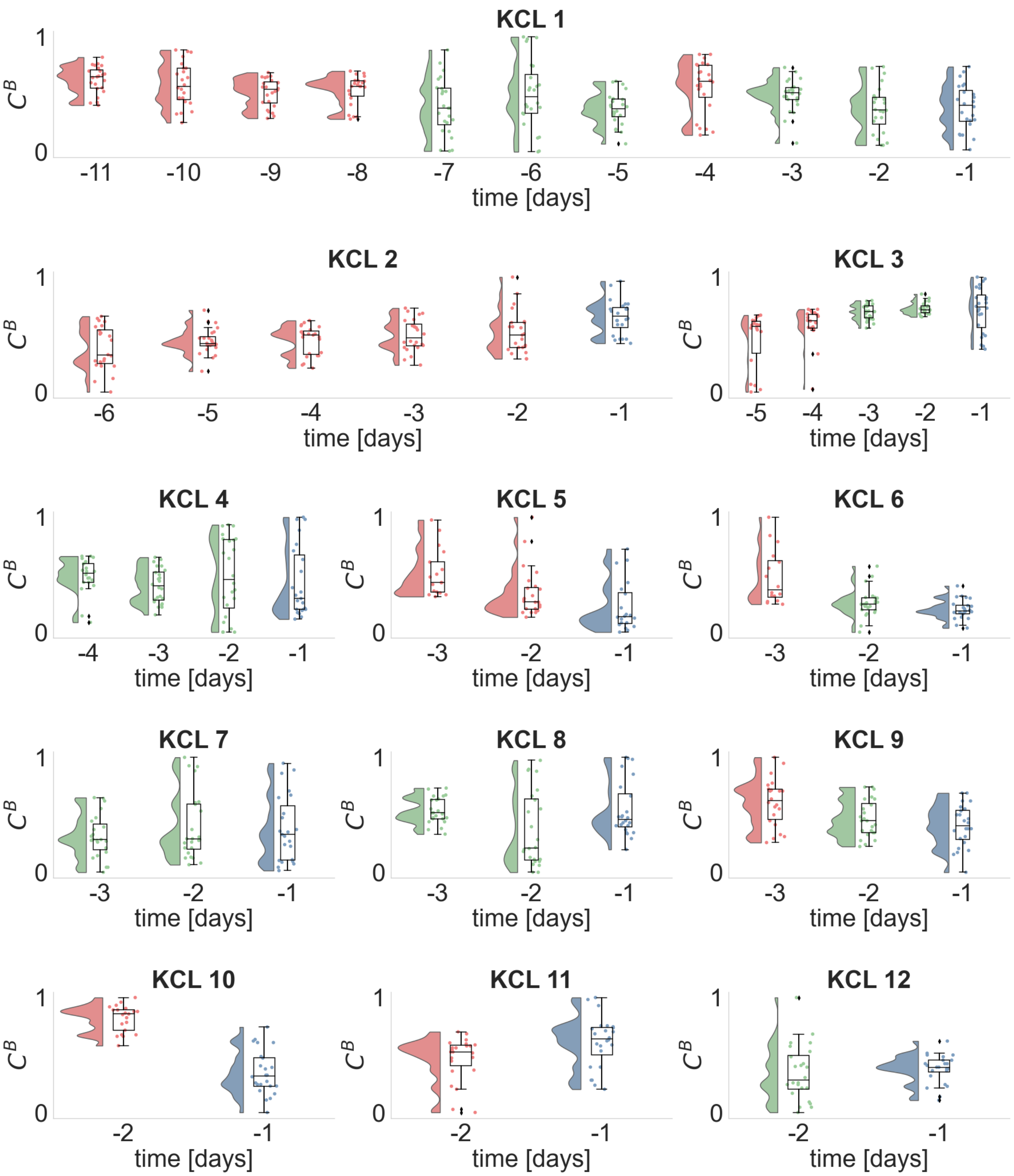


**Figure S13**. Same as Figure S4 for the average betweenness centrality and alpha band.

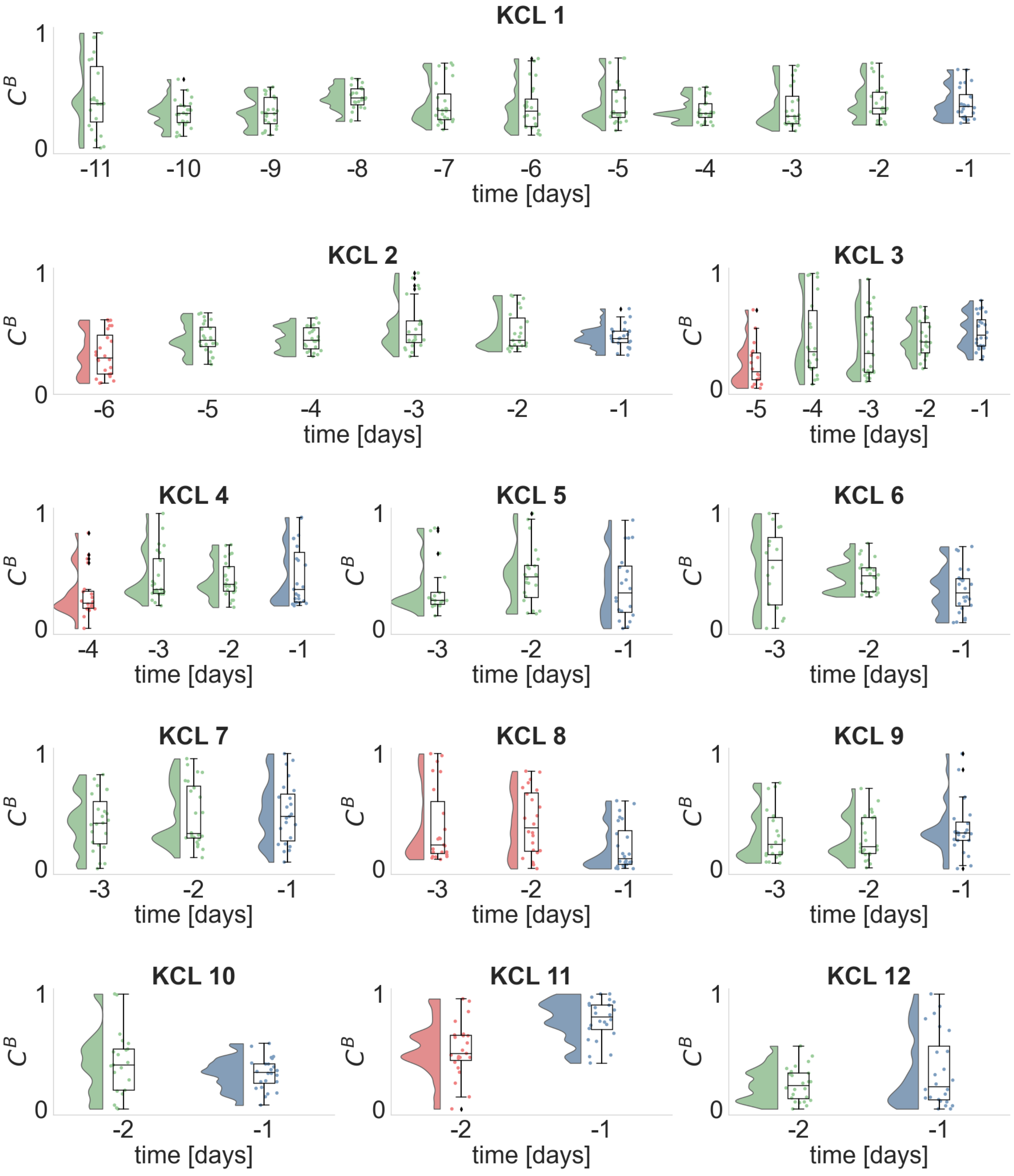


**Figure S14**. Same as Figure S4 for the average betweenness centrality and beta band.

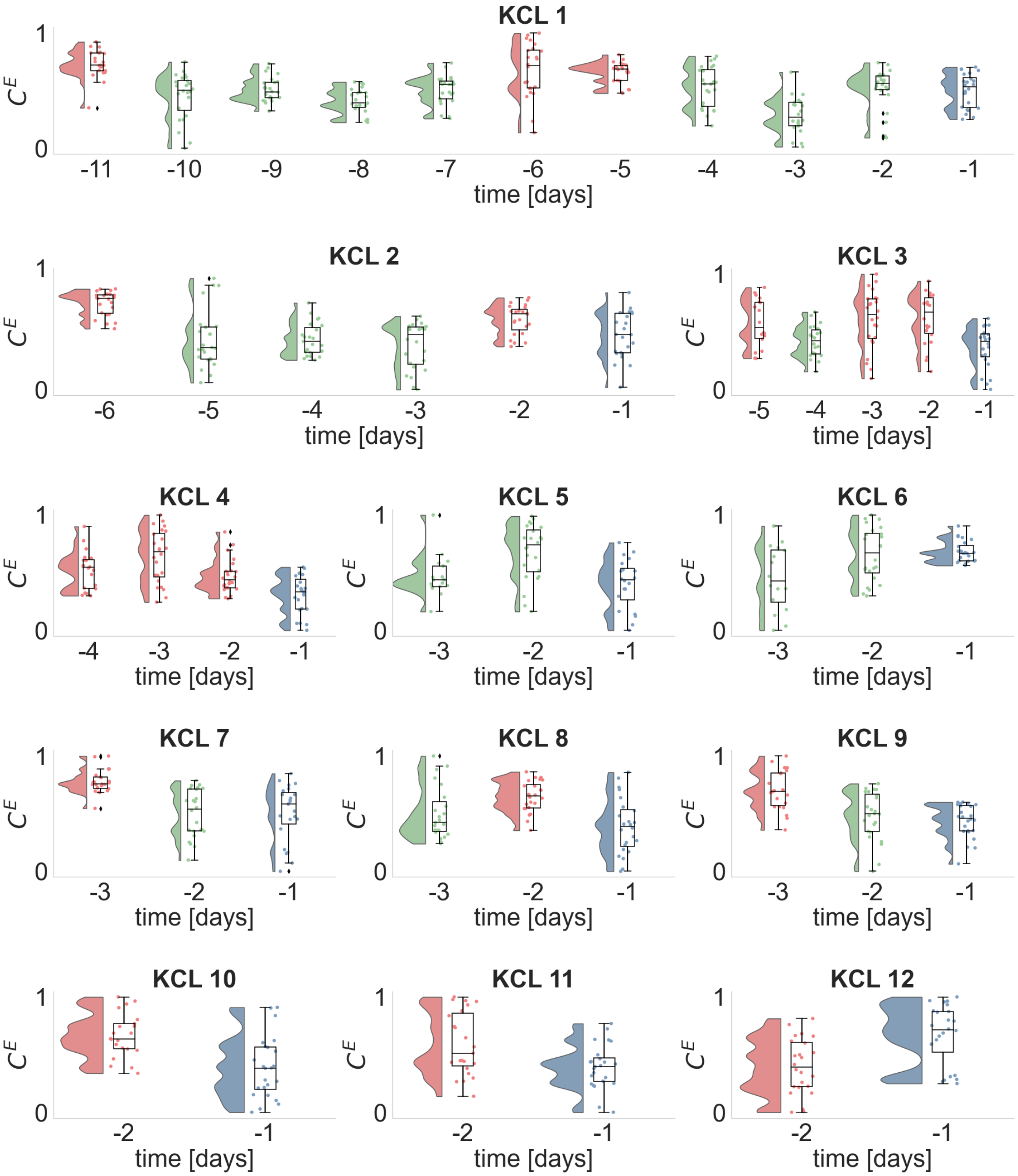


**Figure S15**. Same as Figure S4 for the average eigenvector centrality and delta band.

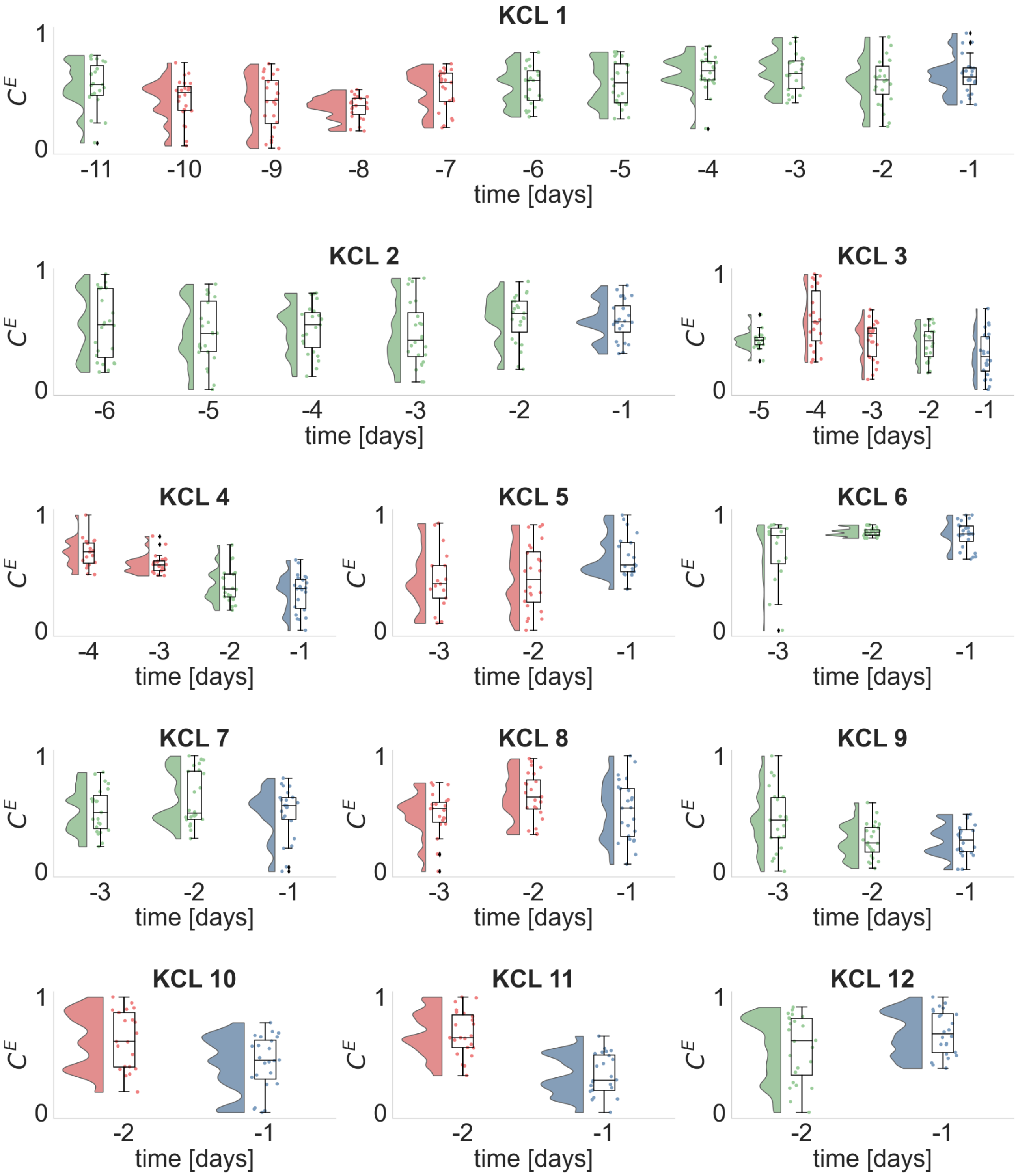


**Figure S16**. Same as Figure S4 for the average eigenvector centrality and theta band.

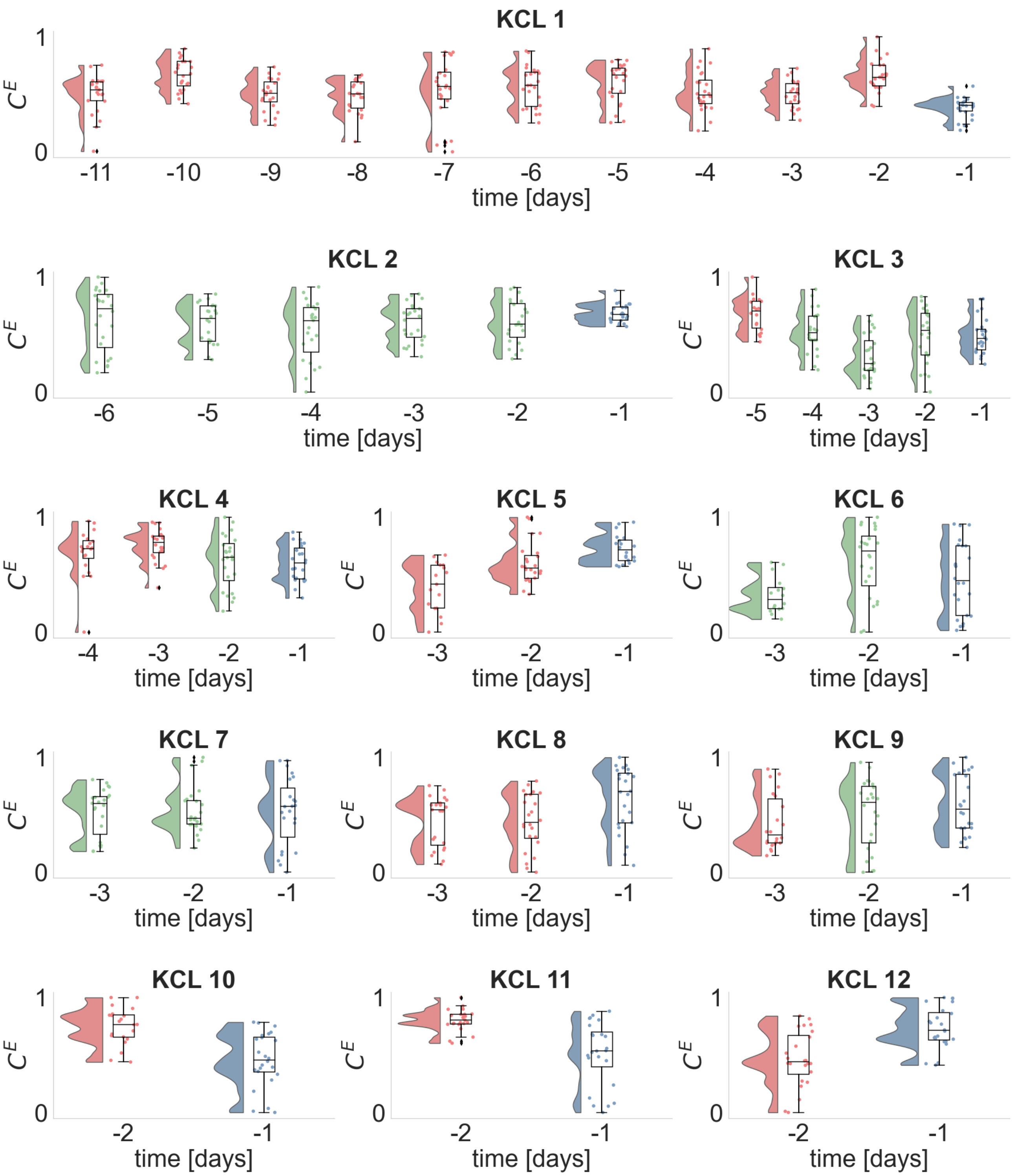


**Figure S17**. Same as Figure S4 for the average eigenvector centrality and alpha band.

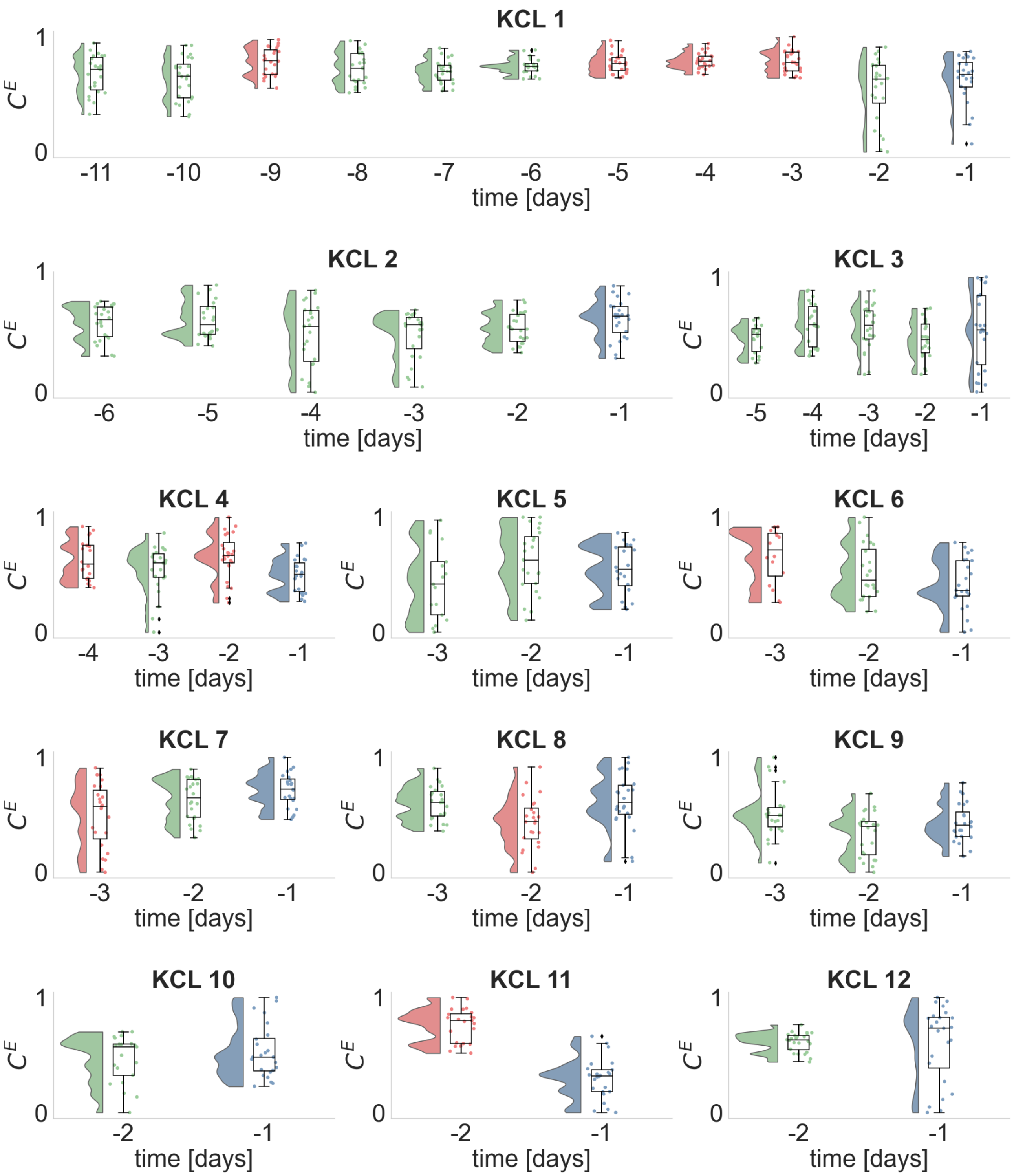


**Figure S18**. Same as Figure S4 for the average eigenvector centrality and beta band.

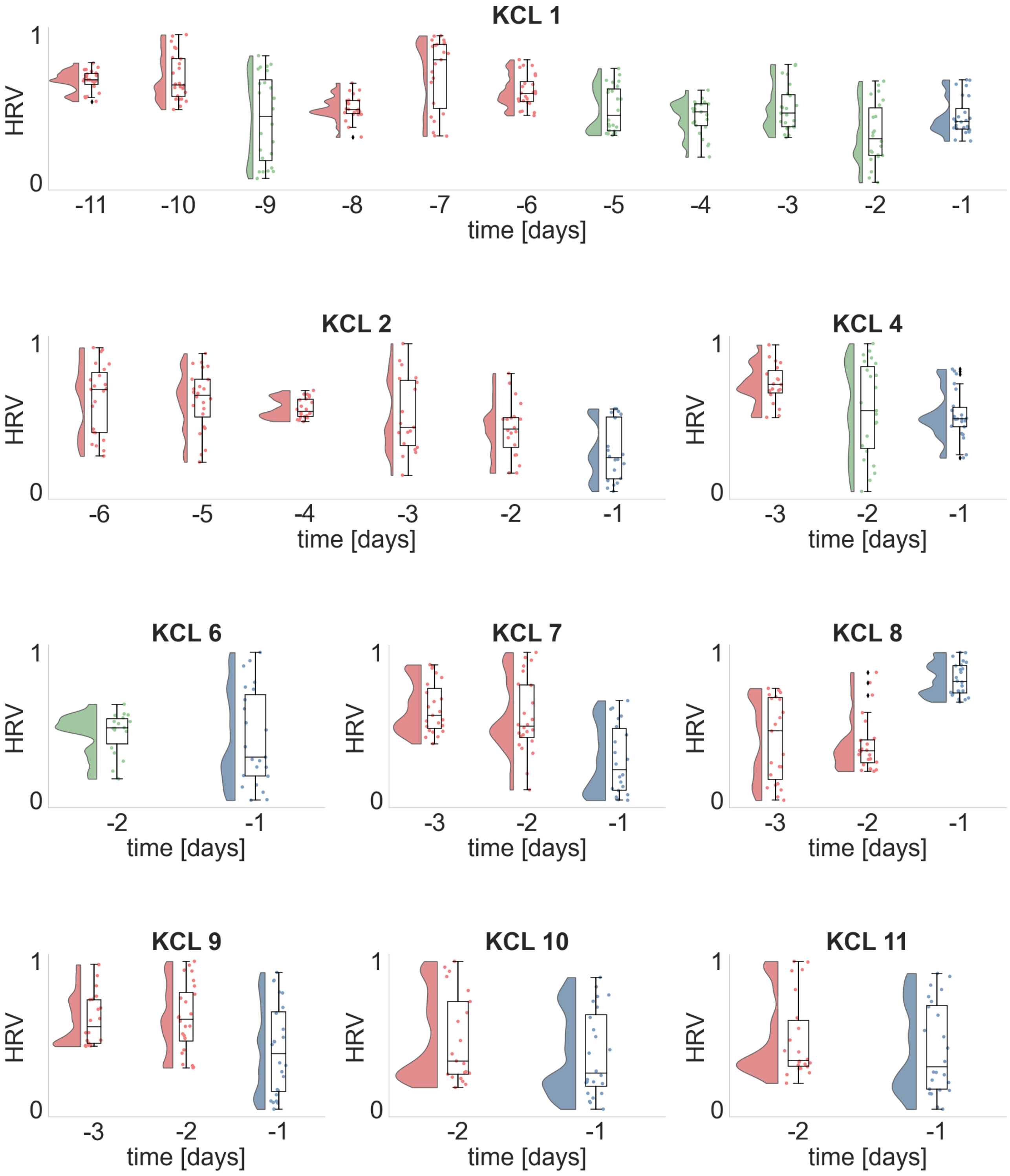


**Figure S19**. Daily distributions of the heart rate variability HRV for each seizure free day prior to seizure occurrence. Dots illustrate the HRV values obtained from each analyzed ECG segment, whilst histograms and boxplots depict their distribution. Horizontal lines in the boxplots indicate the median. The day before the seizure occurrence, i.e., $d_{-1}$ is denoted with blue. Days whose distributions are statistically significantly different (one-sided Wilcoxon rank sum test) from the distribution of day $d_{-1}$ are illustrated in red, otherwise are depicted in green.

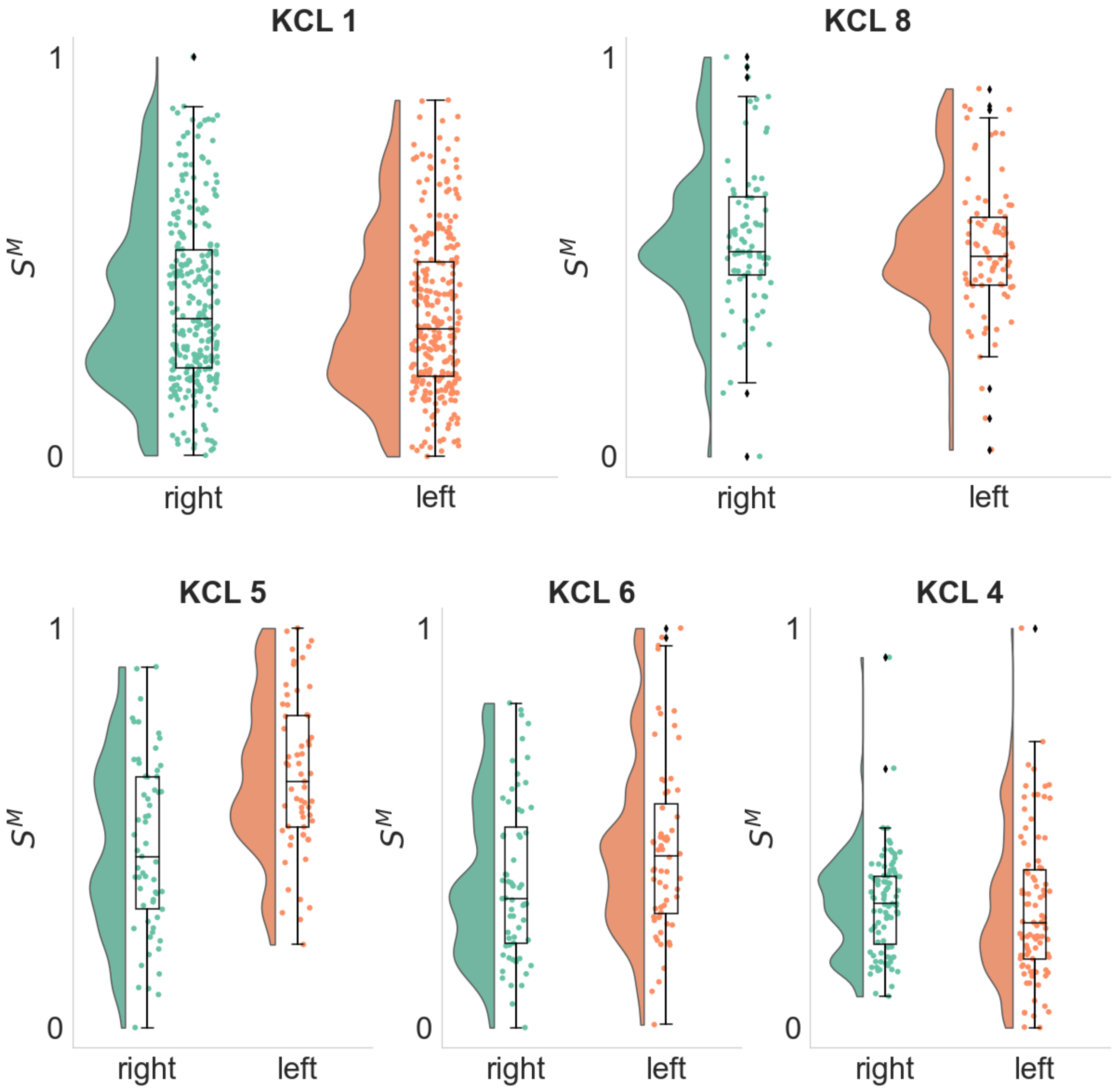


**Figure S20.** Distributions of the Mean Strength $S^M$ of each hemisphere in the alpha frequency band. Each dot depicts the $S^M$ value of the right (green) or left (orange) hemisphere as computed from a single EEG segment. KCL 1 and KCL 2 participants had the seizure focus on the right hemisphere whilst KCL 5, KCL 6 and KCL 4 had the seizure focus on the left hemisphere (p-values= $1.34 \times 10^{-5}$ , only significant for KCL 5, one-sided Wilcoxon rank-sum test).

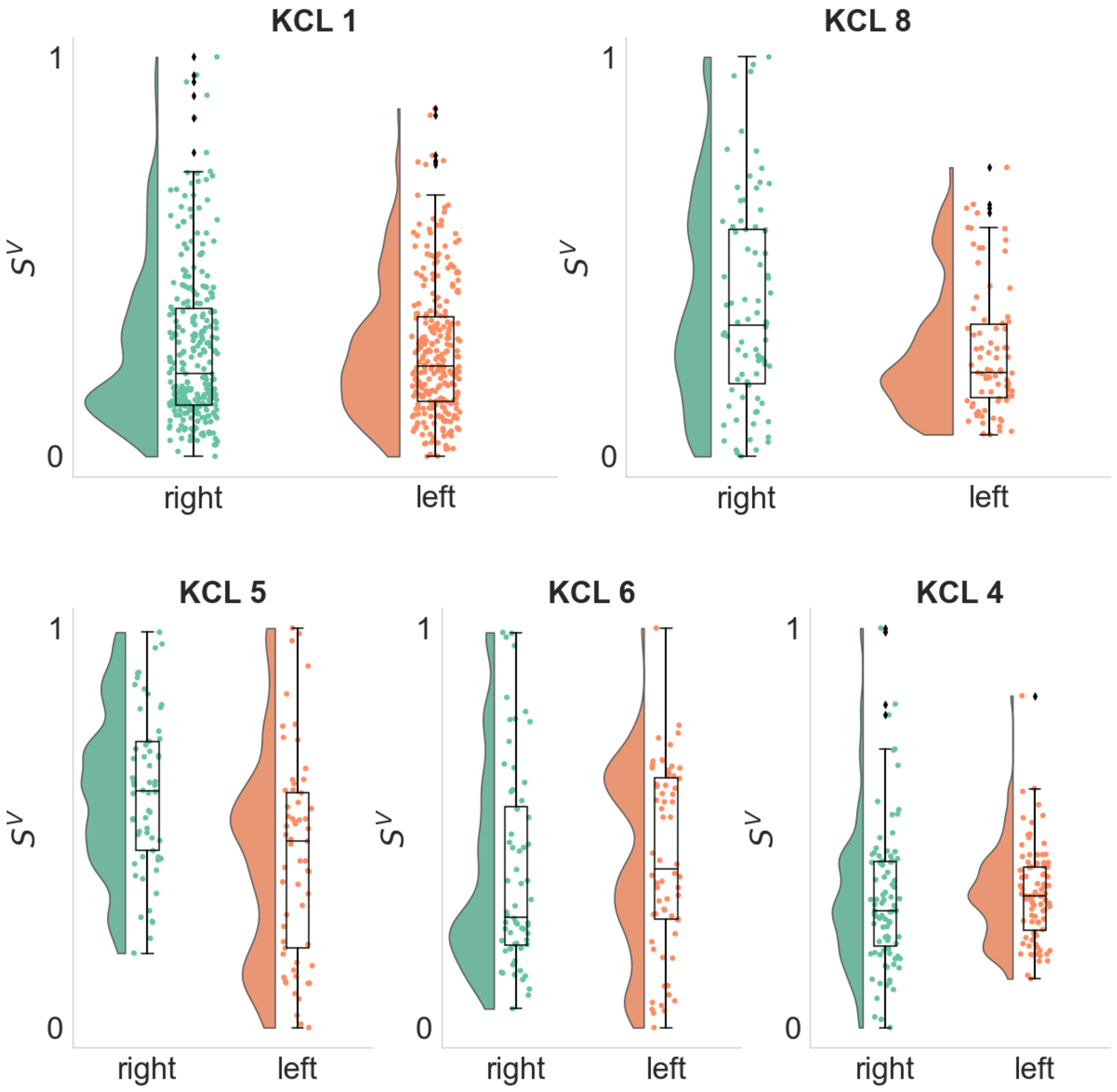


**Figure S21**. Distributions of the Variance of Strength $S^V$ of each hemisphere in the alpha frequency band. Each dot depicts the $S^V$ value of the right (green) or left (orange) hemisphere as computed from a single EEG segment. KCL 1 and KCL 2 participants had the seizure focus on the right hemisphere whilst KCL 5, KCL 6 and KCL 4 had the seizure focus on the left hemisphere (p-values= 0.001 and 0.0001, only significant for KCL 8 and KCL 5, respectively one-sided Wilcoxon rank-sum test).

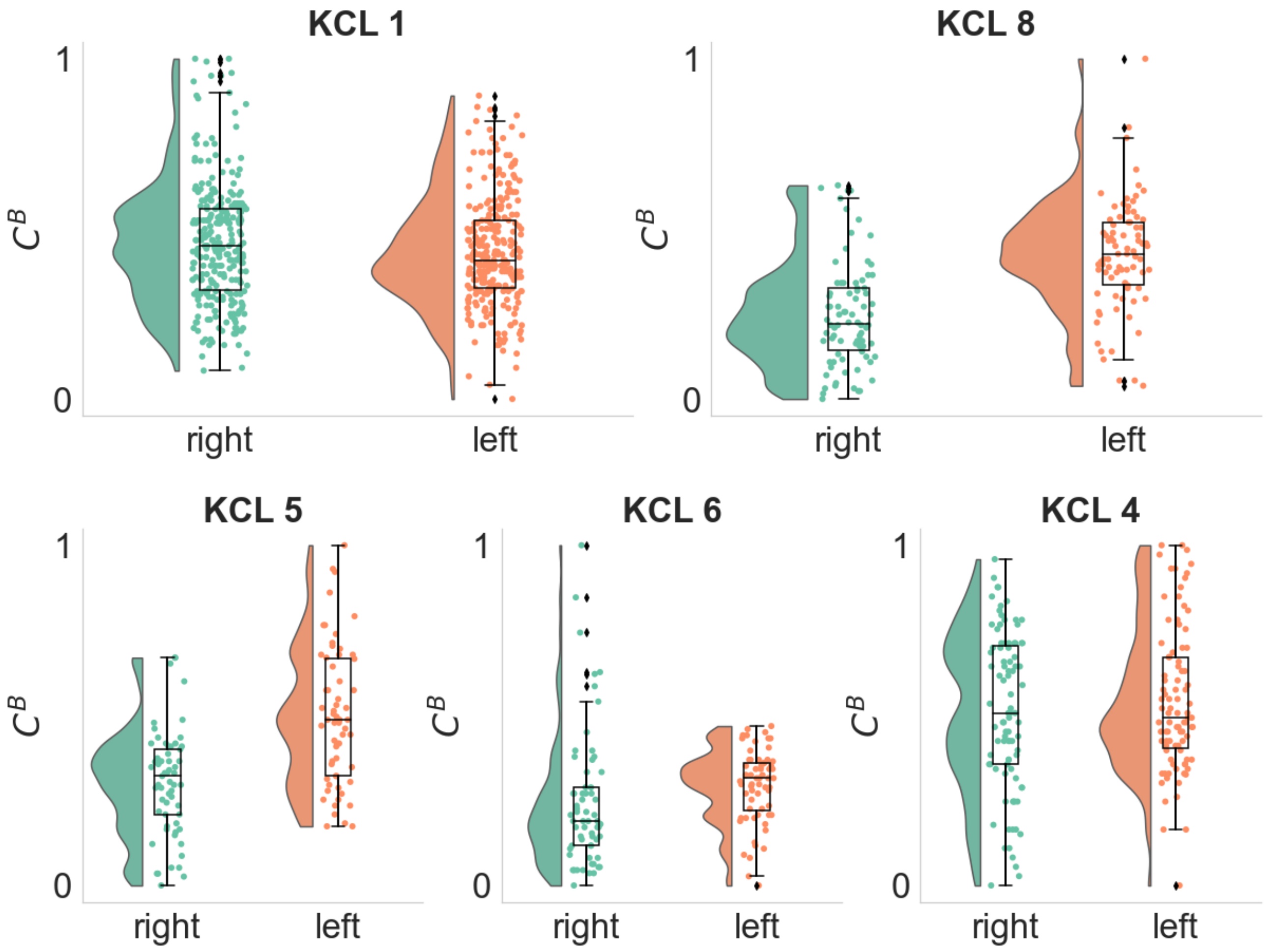


**Figure S22**. Distributions of the Betweenness Centrality $C^B$ of each hemisphere in the alpha frequency band. Each dot depicts the $C^B$ value of the right (green) or left (orange) hemisphere as computed from a single EEG segment. KCL 1 and KCL 2 participants had the seizure focus on the right hemisphere whilst KCL 5, KCL 6 and KCL 4 had the seizure focus on the left hemisphere (p-values= $1.41 \times 10^{-7}$ and 0.0002, only significant for KCL 5 and KCL 6, respectively one-sided Wilcoxon rank-sum test. In KCL 8 there is a statistically significantly difference between the two distributions p-value = $6.24 \times 10^{-11}$, however it is in the opposite direction).
